# Identification and Quantification of Bioactive Compounds Suppressing SARS-CoV-2 Signals in Wastewater-based Epidemiology Surveillance

**DOI:** 10.1101/2022.03.09.22272155

**Authors:** Mohamed Bayati, Hsin-Yeh Hsieh, Shu-Yu Hsu, Chenhui Li, Elizabeth Rogers, Anthony Belenchia, Sally A. Zemmer, Todd Blanc, Cindy LePage, Jessica Klutts, Melissa Reynolds, Elizabeth Semkiw, Hwei-Yiing Johnson, Trevor Foley, Chris G. Wieberg, Jeff Wenzel, Terri Lyddon, Mary LePique, Clayton Rushford, Braxton Salcedo, Kara Young, Madalyn Graham, Reinier Suarez, Anarose Ford, Zhentian Lei, Lloyd Sumner, Brian P. Mooney, Xing Wei, C. Michael Greenlief, Marc C. Johnson, Chung-Ho Lin

## Abstract

Recent SARS-CoV-2 wastewater-based epidemiology (WBE) surveillance have documented a positive correlation between the number of COVID-19 patients in a sewershed and the level of viral genetic material in the wastewater. Efforts have been made to use the wastewater SARS-CoV-2 viral load to predict the infected population within each sewershed using a multivariable regression approach. However, reported clear and sustained variability in SARS-CoV-2 viral load among treatment facilities receiving industrial wastewater have made clinical prediction challenging. Several classes of molecules released by regional industries and manufacturing facilities, particularly the food processing industry, can significantly suppress the SARS-CoV-2 signals in wastewater by breaking down the lipid-bilayer of the membranes. Therefore, a systematic ranking process in conjugation with metabolomic analysis was developed to identify the wastewater treatment facilities exhibiting SARS-CoV-2 suppression and identify and quantify the chemicals suppressing the SARS-COV-2 signals. By ranking the viral load per diagnosed case among the sewersheds, we successfully identified the wastewater treatment facilities in Missouri, USA that exhibit SARS-CoV-2 suppression (significantly lower than 5 × 10^11^ gene copies/reported case) and determined their suppression rates. Through both untargeted global chemical profiling and targeted analysis of wastewater samples, 40 compounds were identified as candidates of SARS-CoV-2 signal suppression. Among these compounds, 14 had higher concentrations in wastewater treatment facilities that exhibited SARS-CoV-2 signal suppression compared to the unsuppressed control facilities. Stepwise regression analyses indicated that 4-nonylphenol, palmitelaidic acid, sodium oleate, and polyethylene glycol dioleate are positively correlated with SARS-CoV-2 signal suppression rates. Suppression activities were further confirmed by incubation studies, and the suppression kinetics for each bioactive compound were determined. According to the results of these experiments, bioactive molecules in wastewater can significantly reduce the stability of SARS-CoV-2 genetic marker signals. Based on the concentrations of these chemical suppressors, a correction factor could be developed to achieve more reliable and unbiased surveillance results for wastewater treatment facilities that receive wastewater from similar industries.

## 1. Introduction

Coronaviridae (Coronavirus) is a family of positive sense single stranded RNA viruses, responsible for various severe respiratory infections [1, 2]. This family contains over 30 kinds of viruses and has a genome of approximately 30 Kb, the largest reported genome of all RNA viruses [3, 4]. In the past 17 years, there have been three major outbreaks caused by human coronaviruses, including the severe acute respiratory syndrome coronavirus (SARS-CoV) that occurred in China in 2003 and affected 26 countries [5, 6]. In 2012, the outbreak of the Middle East Respiratory Syndrome Coronavirus (MERS-CoV) [5, 7] affected 27 countries with over 2,400 cases [8]. Recently, the ongoing Coronavirus Disease 2019 (COVID-19) pandemic caused by Acute Respiratory Syndrome Coronavirus 2 (SARS-CoV-2) that emerged in Wuhan, China, has affected the global community and individual daily function [9–11]. Recent studies have revealed that both SARS-CoV-2 and SARS-CoV can recognize and bind to the angiotensin-converting enzyme 2 (ACE2) on the cell surface. Between the two viruses, subtle differences in the amino acid sequence in addition to conformation of the S protein in SARS-CoV-2 contribute to a significantly stronger affinity of SARS-CoV-2 to ACE2 [12, 13]. ACE2 is not only highly expressed in lungs, but also in the gastrointestinal tract, including the small intestine and colon [14].

Wastewater-based epidemiology (WBE) has been used as a surveillance tool for population-wide infectious diseases, featuring a proven track record for hepatitis A and polio [15]. Different studies in the United States, the Netherlands, Italy, and elsewhere have detected the presence of SARS-CoV-2 in domestic sewage and have found a positive relationship between the amount of viral material in sewage and the number of reported COVID-19 cases in the area that collects and treats wastewater for a community, called a “sewershed”[16–19]. Although a majority of the SARS-CoV-2 viral loads in wastewater are introduced through the gastrointestinal tract, SARS-CoV-2 can also be introduced into wastewater (domestic and hospital) through several other sources, such as sputum, handwashing, and vomit [20–22]. However, the main source of SARS-CoV-2 viral loads to wastewater that has been reported is feces containing viral RNA shed by infected people [23–26].

Due to the documented positive correlation between the number of COVID-19 patients in a sewershed and the level of viral genetic material in the wastewater in recent SARS-CoV-2 WBE studies [27, 28], efforts have been made to use the wastewater SARS-CoV-2 viral load to predict the infected population for each sewershed using a multivariable regression approach. However, reported clear and sustained variability among treatment facilities have made clinical prediction challenging. Specifically, wastewater at some facilities consistently exhibits higher genetic material per diagnosed patient, indicating a likely underestimate in the number of COVID-19 patients, while wastewater from other facilities has much lower levels of the genetic material per diagnosed case, suggesting suppression of the genetic material from the sewershed. Since it is quite common that wastewater treatment facilities receive some input from industries, several classes of molecules released by regional industries and manufacturing facilities, particularly the food processing industry, could significantly suppress SARS-CoV-2 signals in wastewater by breaking down the lipid-bilayer of the membranes [29–32].

The active ingredients in detergents, surface-active agents (surfactants), emulsifiers, and disinfection products (e.g., pyrrolidones, sodium dodecylbenzinesulfonate, sodium xylenesulfonate, polyethylene glycol, sodium stearate and cocamidopropyl betaine), as well as bioconugate and cross-linking agents (e.g., ethylenediaminetetraacetic acid) are commonly found in industrial wastewater[33–37]. Among these chemicals, surfactants are one of the main compounds that can be exist in wastewater [41]. The surfactants consist of two major functional groups: one is hydrophilic (lipophobic) and the other is non-polar hydrophobic (lipophilic) [35]. Generally, the two functional groups are referred as head and tail, respectively. The surface-active agents are usually classified based on the charge of the head, including anionic, cationic, non-ionic and zwitterionic compounds. Approximately 65% of the total world production of surfactants corresponds to the compounds classified as anionic surfactants [35, 38]. Surfactants are mainly used in surface cleaners, household detergents, shampoos, dishwashing liquids, cosmetics, and laundry detergents [39]. Moreover, different varieties of surfactants are used as starting materials in the production of pigments, catalysts, dyes, pesticides, pharmaceuticals, and plasticizers [40]. These compounds could significantly reduce the stability of SARS-COV-2 genetic marker signals in wastewater by breaking down the lipid bilayer of SARS-COV-2. Therefore, for facilities that receive wastewater from industries, a correction factor based on the concentrations of such bioactive molecules is needed to achieve more reliable and unbiased surveillance results.

As a result of recent advancements in mass spectrometry, metabolomics algorithm, computational capacity, and mass spectral reference databases, untargeted metabolomics has been widely applied to identify bioactive molecules in the complex and organic-rich matrices. Untargeted metabolomics is the global profiling of small molecules in a system without any bias. Although several analytical techniques can be employed to perform untargeted metabolomics, liquid chromatography coupled with high-resolution mass spectrometry (LC/HRMS) has been frequently used because of the large number of molecules that can be evaluated in a single analysis [42]. For example, ten to thousands of features (a feature is defined as an ion with a distinctive m/z and retention time) can be detected by high resolution LC/HRMS in one extract.

In general, the main purpose of untargeted metabolomics is to determine which of these features is dysregulated (upregulated and downregulated) between different sample groups or treatments. Due to the complexity and the number of features in a dataset, it is challenging to accomplish this comparison manually [43]. Several software programs for automated processing of LC/HRMS data have been developed over the past decade. However, most of these programs have restrictions that limit their utility and applicability to different instrumentation. One widely applicable program for processing LC/HRMS data is XCMS Online, a web-based platform that contains all of the tools necessary for the entire untargeted metabolomic workflow, including signal detection, peak alignment, retention time correction calculations, raw data processing, statistical analysis, and metabolite assignment [42–44]. An untargeted metabolomic profiling approach that utilizes a comprehensive program like XCMS Online is well-suited to the identification of candidate compounds that suppress the SARS-CoV-2 genetic signal in complex wastewater matrices.

The objectives of this study are to 1) identify the wastewater treatment facilities in Missouri, USA that exhibit SARS-CoV-2 suppression and determine their suppression rates, 2) identify possible active compounds suppressing the SARS-CoV-2 genetic signal through a combination of stepwise regression and metabolomic profiling approaches, 3) confirm and quantify the identified bioactive molecules using targeted analysis, and 4) validate the suppression activities through incubation studies.

## 2. Materials and Methods

### 2.1. Materials

High performance liquid chromatography (HPLC) grade methanol (MeOH), acetonitrile (ACN), and formic acid (FA) were purchased from Sigma-Aldrich (St. Louis, MO, USA). HPLC grade ammonium acetate was purchased from Fisher Scientific (Pittsburgh, PA, USA). Analytical standards were purchased from Sigma-Aldrich unless otherwise mentioned. The TaqPath™ 1-Step RT-qPCR Master Mix and TaqMan Probes were purchased from Thermo Fisher Scientific. The primers and probes used in the qPCR assay were purchased from Integrated DNA Technologies, Inc. (Coralville, IA, USA).

### 2.2. Wastewater Sample Collection

From July-December 2020, more than fifty-seven wastewater treatment facilities across the state of Missouri, USA were monitored weekly for SARS-CoV-2. The wastewater samples were gathered from the influent of the wastewater treatment facilities (i.e., prior to primary treatment) (**Table S1**). Once per week, triplicate 50 mL subsamples were collected in polypropylene centrifuge tubes from the 24-hour composite wastewater samples. Subsamples kept chilled (between 0 and 3 °C) during transportation to the laboratory at the University of Missouri in Columbia. All the samples were stored at -20 °C until they were analyzed.

**Table 1.**
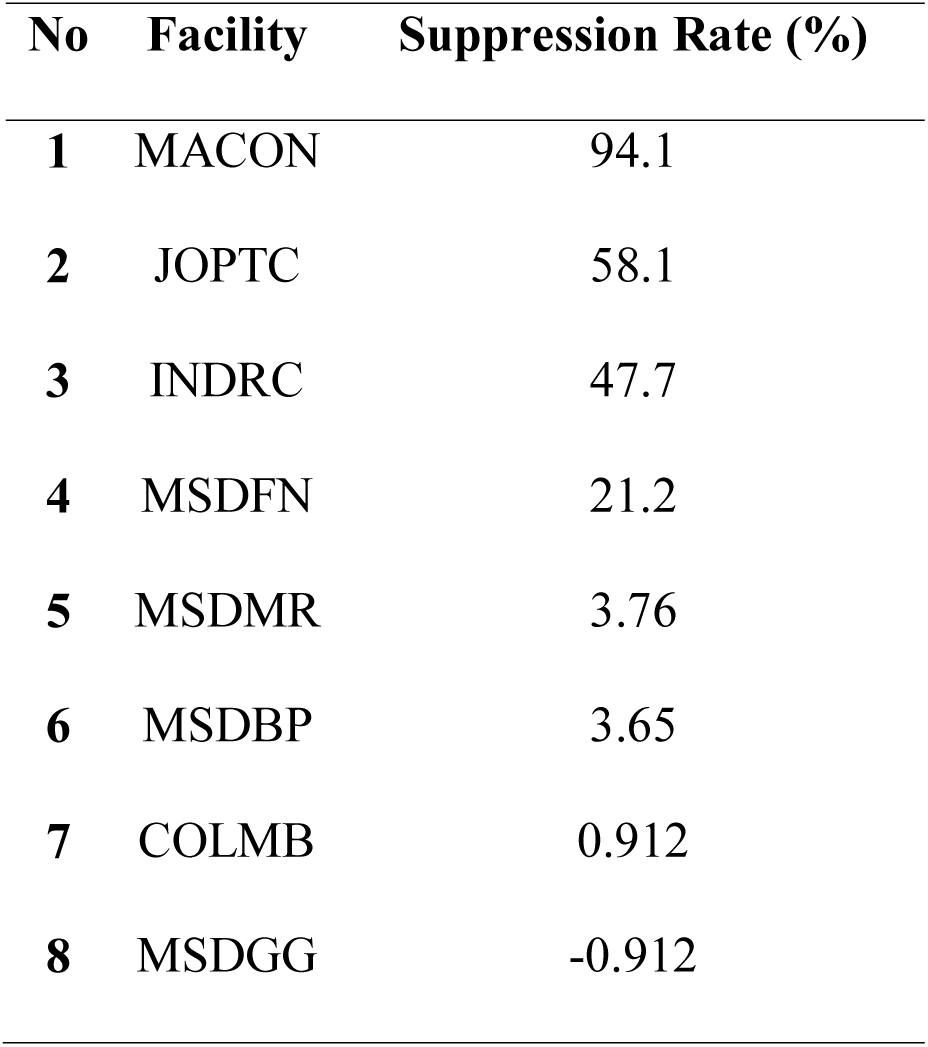
SARS-CoV-2 gene suppression rates for the facilities included in this study.

| No | Facility | Suppression Rate (%) |
| --- | --- | --- |
| 1 | MACON | 94.1 |
| 2 | JOPTC | 58.1 |
| 3 | INDRC | 47.7 |
| 4 | MSDFN | 21.2 |
| 5 | MSDMR | 3.76 |
| 6 | MSDBP | 3.65 |
| 7 | COLMB | 0.912 |
| 8 | MSDGG | -0.912 |

### 2.3. Quantification of SARS-CoV-2 in Wastewater

#### 2.3.1. RNA Extraction from Wastewater Samples

Fifty mL of wastewater from the catchment were filtered through a 0.22 µm filter (Millipore cat# SCGPOO525). Thirty-six mL of filtered wastewater were mixed with 12 mL of 50% (W/V) polyethylene glycol (PEG, Research Products International, cat# P48080) and 1.2 M NaCl, followed by incubation for 1 h at 4°C. Samples were further centrifuged at 12,000 rpm for 2 h. RNA was extracted from the pellet using Qiagen Viral RNA extraction kit following the manufacturer’s instructions after the supernatant was removed. RNA was eluted in a final volume of 60 µL. The samples were stored at -20°C if not processed immediately.

#### 2.3.2. Plasmid Standard

A plasmid carrying a puromycin resistance (*puro)* gene fragment (5’ ATGACAGAGTATAAGCCAACCGTCCGGCTCGCAACGAGAGACGATGTCCCGAGGGCAGTGCGCACGCTCGCCGCGGCCTTTGCGGACTACCCTGCAACAAGACACACTGTGGATCCCGATCGCCACATAGAGCGCGTGACTGAGCTGCAAGAACTGTTCCTTACCAGGGTGGGTCTCGATATCGGTAAGGTTTGGGTCGCCGACGACGGAGCGGCAGTGGCAGTCTGGACCACTCCTGAGAGCGTAGAAGCAGGCGCAGTGTTTGCAGAAATTGGCCCTAGAATGGCCGAATTGTCCGGTAGCCGGCTCGCTGCTCAGCAGCAGATGGAAGGCCTGCTCGCACCTCACAGACCCAAAGAACCCGCGTGGTTCCTGGCGACAGTGGGAGTCAGTCCAGACCATCAGGGCAAAGGTCTCGGCTCAGCAGTTGTACTGCCTGGGGTAGAGGCCGCAGAAAGGGCAGGGGTGCCGGCCTTCCTGGAAACATCTGCACCCAGAAACTTGCCTTTCTACGAGAGGCTGGGATTCACCGTTACCGCCGACGTGGAGGTGCCCGAAGG ACCGCGCACTTGGTGCATGACGAGAAAGCCCGGGGCTTGA 3’) along with a N gene fragment were constructed, purified from *Escherichia coli*, and used as standards for the RT-qPCR assay. The primer pair (COVID19-N 5p: 5’ ATGTCTGATAATGGACCCCAAAATCAGCG 3; COVID19-N 3p: 5’ TTAGGCCTGAGTTGAGTCAGCACTGC 3’) was used to amplify the N ORF fragment from IDT’s 2019-nCoV_N_Positive Control plasmid and the N ORF fragments were infused using an InFusion kit (Takara). A standard curve was constructed at concentrations of 200,000 through 2 gene copies µL^-1^ and utilized to determine the copy number of the target *puro* gene in the spiked wastewater samples.

#### 2.3.3. Quantitative RT-qPCR Assay

The TaqMan probe (FAM-5’ ACCCCGCATTACGTTTGGTGGACC 3’ BHQ1) and the primer pair (2019-nCoV_N1-F: 5’ GACCCCAAAATCAGCGAAAT 3’; 2019-nCoV_N1-R: 5’ TCTGGTTACTGCCAGTTGAATCTG 3’) for N1 detection, and The TaqMan probe (FAM 5’ ACAATTTGCCCCCAGCGCTTCAG 3’ BHQ1) and the primer pair (2019-nCoV_N2-F: 5’ TTACAAACATTGGCCGCAAA 3’; 2019-nCoV_N2-R: 5’ GCGCGACATTCCGAAGAA 3’) for N2 detection were purchased from Integrated DNA Technologies (IDT), based on the CDC 2019-nCoV Real-Time RT-PCR Diagnostic Panel (Acceptable Alternative Primer and Probe Sets) https://www.cdc.gov/coronavirus/2019-ncov/downloads/List-of-Acceptable-Commercial-Primers-Probes.pdf. The TaqMan probe (VIC-5’ CGGTAAGGTTTGGGTCGCCGAC 3’-QSY) and the primer pair (*puro* Forward: 5’ CCCGATCGCCACATAGAGC 3’; *puro* Reverse: 5’ CCATTCTAGGGCCAATTTCTGC 3’) were designed and used to target the *puro* RNA described above. A plasmid (described above) carrying a unique *puro* resistance gene fragment along with a N gene fragment was constructed, purified from *Escherichia coli*, and used as standards for the RT-qPCR assay to ensure an equal molar ratio of puro and N gene detection. A standard curve was constructed at concentrations of 200,000 through 2 gene copies μL^-1^ and utilized to determine the copy number of the target *puro* gene in the spiked wastewater samples.

Final RT-qPCR one step mixtures consisted of 5 µL TaqPath 1-step RT-qPCR Master Mix (Thermo Fisher), 500 nM of each primer, 125 nM of each of TaqMan probes, 5 µL of wastewater RNA extract and RNase/DNase-free water to reach a final volume of 20 µL. All RT-qPCR assays were performed in duplicate using a 7500 Fast real-time qPCR machine (Applied Biosystems). The reactions were initiated with 1 cycle of uracil-DNA glycosylase (UNG) incubation at 25 C for 2 min and then 1 cycle of reverse transcription at 50 C for 15 min, followed by 1 cycle of activation of DNA polymerase at 95 C for 15 min, C for 2 min and then 45 cycles of 95 C for 3 sec for DNA denaturation and 55 C for 30 sec for anneal and extension. The data was collected at the step of 55 C extension.

### 2.4. Determination of Average Wastewater SARS-CoV-2 Viral Load for Each Reported Patient to Identify the Facilities Exhibiting Suppression

In order to predict the average SARS-CoV-2 gene copies produced by each patient contributing to the sewershed as the benchmark for assessing the suppression rate for each facility. Fifty-seven facilities were monitored from July 6, 2020, to December 7, 2020. Wastewater samples were collected in triplicate from each facility once a week during that period. Flow rates information was collected by the wastewater operators, while the number of cases reported for each sewershed was provided by the Department of Health and Senior Services (DHSS). To establish the relationship between SARS-CoV-2 viral load and case number, the total viral loads were calculated according to Eq. (1):

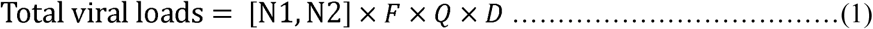

where [N1, N2] (copies/µL) is the average SARS-CoV-2 concentration in the wastewater samples, determined by RT-qPCR. *F* is the extraction factor (350), that converts the units from copies/µL to copies/L. *Q* is the flow rate (L/day), and *D* is the number of days (161 days). The average viral load per diagnosed case was calculated by developing a regression relationship between the viral load and diagnosed case numbers.

### 2.5. Identifying the Facilities for Chemical Analysis

The facilities consistently showing low viral load per diagnosed case which are deviated from the established correlation between viral load and reported cases, suggests suppression of the viral genetic material from the sewershed, were identified. Thus, the viral load per diagnosed case for all the 57 tested facilities were ranked according to their standardized suppression rates.

To develop the relationship between suppression rates and the concentrations of each identified molecule, the facilities representing a gradient of suppression rates, including no suppression, moderately suppression and severely suppression, were selected for further chemical analysis. The chemical analysis in combination of the stepwise regression analysis were integrated to help identify the bioactive compounds that suppressed the SARS-CoV-2 signals.

### 2.6. Sample Preparation for Chemical Profiling and Targeted Analysis

The wastewater samples collected in 50 mL polypropylene centrifuge tubes were vortexed (Vortex Genie 2, Fisher, NY, USA) for 10 sec before being transferred to smaller tubes. Then, 1.8 mL of the wastewater was transferred to 2 mL microcentrifuge tubes and centrifuged (Eppendorf 5415D, Hamburg, Germany) at 12,000 rpm for 15 min. After centrifugation, 1.5 mL of the wastewater supernatant and 1.5 mL MeOH were mixed in 5 mL glass tubes. The mixture was vortexed for 10 sec and 1.5 mL was filtered through 0.2 µm syringe filter (Acrodisc with PTFE membrane, Waters, MA, USA). Extracts were stored at -20 °C until analysis with the high-performance liquid chromatography-tandem mass spectrometry (LC-MS/MS).

### 2.7. Untargeted Metabolomics Global Chemical Profiling Analyses

Ultra-High Performance Liquid Chromatography (UHPLC) system coupled to a maXis impact quadrupole-time-of-flight high-resolution mass spectrometer (Q-TOF) (Bruker Co., Billerica, MA, United States) was used to analyze the wastewater extracts. The system was operated in either negative or positive electrospray ionization modes with the nebulization gas pressure at 43.5 psi, dry gas of 12 L/min, dry temperature of 250 C and a capillary voltage of 4000 V. The wastewater samples from 8 different locations were separated using Waters Acquity UHPLC BEH C18 column (2.1 × 0 mm, 1.7 mm particles size) at 60 C. The solvent system was 0.1% formic acid (FA) in water (A) and 100% acetonitrile (B). The gradient elution used started with a linear gradient of 95%: 5–30%: 70% (eluents A: B) in 30 min. Subsequently, the separation was followed by a linear wash gradient as follows 70–95% B, 95% B, 95–5% B, and 5% B at 30–33 min, 33– 35 min, 35–36 min, and 37–40 min, respectively. The flow rate was 0.56 mL/min. Mass spectral data were collected automatically using a scan range from 100 to 1,500 m/z and auto calibrated using sodium formate after data acquisition. Each wastewater sample and methanol blank (control) were analyzed in triplicate.

To identify the molecules of interest that exhibited statistically significant differences in relative intensities among the wastewater treatment facilities, the CDF files obtained from UHPLC-MS analysis were uploaded and processed using XCMS Online (xcmsonline.scripps.edu). XCMS is a cloud-based informatics platform that can process and visualize mass-spectrometry-based untargeted metabolomic data and perform statistical analysis [22][23]. The data process includes spectra extraction, peak grouping, peak detection, and retention time alignment. Pair comparisons were used for two groups (i.e., wastewater extracts and MeOH control blanks). The XCMS data were processed using the following parameters: pairwise jobs between each wastewater extract and the control (methanol) were conducted in centWave mode for feature detection (minimum peak width = 5 s, 1 m/z = 10 ppm, and maximum peak width = 20 s), an obiwarp method was selected for retention time correction (profStep = 1), chromatogram alignment was set as minfrac = 0.5, bw = 5, mzwid = 0.015, max = 100, minsamp = 1, and adducts were optimized for UPLC/Bruker Q-TOF in both positive and negative ESI mode. An unpaired parametric Welch t-test was used for the statistical analysis. Metabolites of significant features (p < 0.001 and intensity ≥10,000) were putatively identified by the integration of the METLIN database with XCMS Online. To further characterize and visualize the differences in profiles of compounds among different facilities, partial least squares-discriminant analysis (PLS-DA) was performed and heatmap was generated via the web-based tool MetaboAnalyst (Wishart Research Group, University of Alberta, Alberta, Canada) [47].

### 2.8. Targeted Analyses for Confirmation and Quantification

The compounds identified through untargeted analysis were quantified using liquid chromatography-tandem mass spectrometry (LC-MS/MS). The LC-MS/MS analyses were performed using an HPLC system (Water Alliance 2695, Water Co., Milford, MA, United States) coupled with a Waters Acquity TQ triple quadrupole mass spectrometer operated in negative and positive electrospray ionization modes with the nebulization gas pressure at 43.5 psi, dry gas of 12 L/min, dry temperature of 250 C and a capillary voltage of 1500 V. Compounds in the wastewater extracts (30 µL volume per injection) were separated using a Phenomenex Kinetex C18 reverse-phase column (100 × 4.6 mm; 2.6 mm particle size, Torrance, 4 CA, United States) at 40 C. The mobile phases were 0.1% formic acid and 10 mM ammonium acetate in water (A) and 100% acetonitrile (B). The elution gradient used was 2% B (0-0.5 min), 2–80% B (0.5–7 min), 80–98% B (7.0–9.0 min), 2% B (9.0–15.0 min) at a flow rate of 0.5 mL/min. MS detection was performed by MS/MS using the multiple reaction monitoring (MRM) mode. Waters IntelliStart optimization software was used to optimize collision and ionization energy, MRM and SIR (single ion recording) transition ions (molecular and product ions), capillary and cone voltage, and desolvation gas flow. Waters Empower 3 software was used to analyze data. Concentrations of the compounds found in wastewater extracts were determined based on a calibration curve for each analyte generated using standards of these compounds (purity > 95%, Sigma-Aldrich) at 8 concentrations (0.01, 0.05, 0.1, 0.5, 1.25, 2.5, 5, 10 ppm) in triplicate. The limit of detection (LOD) and limit of quantification (LOQ) were calculated to assess the sensitivity of the analytical method. For each compound, the signal-to-noise ratios of three and ten were employed to calculate LOD and LOQ, respectively.

The compounds that could not be ionized or detected by the Waters Acquity TQ triple quadrupole, including 4-octylphenol, sodium tetradecyl sulfate, diethylene glycol, netilmicin and dicyclopentadiene, were quantified by a Waters Xevo TQ-S triple quadrupole mass spectrometer coupled to UHPLC system. A symmetry C18 column (2.1×100mm, 3.5µm, WAT058965) was used and compounds separated by gradient delivery (0.5 mL/min) of solvent. Initial conditions were 95%A and 5%C (Solvent A: 0.1% FA, 2mM ammonium acetate, in water; solvent B: acetonitrile with 0.1% FA; solvent C:0.1% FA, 2mM ammonium acetate, in methanol), which ramped to 30% B and 70% C over 3 min, and held at 30%B and 70%C over 3 min, followed by going back to the initial composition within 0.1 min, and being held at the initial conditions for 0.9 min. The total run time was 7 min. The column was heated to 30 °C and the samples were cooled to 20 °C in the autosampler.

### 2.9 Stepwise-Regression Analysis for Identifying the Molecules Suppressing SARS-CoV-2 Signals in Wastewater

Stepwise linear regression models and least absolute shrinkage and selection operator (LASSO) regression models were utilized to identify the compounds that are positively correlated with the SARS-CoV-2 suppression rates. In all models, chemical signal intensities quantified by UHPLC-MS in positive or negative ion mode were the predictor variable and the viral suppression rate at selected WWTPs facilities was the response variable.

Four different statistical approaches were used to determine the positive correlation between the relative intensities of the compounds and suppression rate. The four approaches included: forward stepwise regression, backward stepwise regression, best subset linear regression, and LASSO. The regsubsets() function in the R package leaps (https://cran.r-project.org/web/packages/leaps/leaps.pdf) were utilized for forward, backward and best subset stepwise regression models. The forward selection began with a model without any predictor variables. The predictors were added to the model one by one until all of them were in the model. Conversely, backward selection began with the model with all predictors, followed by leaving one out at a time until no predictor was in the model [48]. The best subset regression model selected the subset model from all combinations of predictors based on the goodness-of-fit criteria [49]. In the end, the subset model with the highest adjusted R^2^ out of all tree approaches was chosen. The linear regression model was then fitted with the chosen predictors, and the coefficients were examined.

To avoid overfitting the model, LASSO regression model was also used to examine the predictors. The glmnet() function in the R package glmnet package (https://cran.r-project.org/web/packages/glmnet/glmnet.pdf) was used to build the model. The shrinkage penalty (λ) was determined by cross validation. The coefficients of insignificant predictors with λ were shrunk to zero [50].

### 2.10 Suppression Study

The suppression experiments were carried out to investigate the effect of the identified molecules on SARS-CoV-2 genetic materials in the wastewater. Stock solutions of each identified compound were prepared with commercially available standards in 100% methanol at a concentration of 10,000 mg/L. A 20 mL wastewater sample with verified high SARS-CoV-2 concentrations was mixed with 20 mL ultrapure water (MilliQ system, 18.2 mΩ.cm at 25 Synergy® Water Purification System, MA, USA). The mixture was stirred gently for 5 min and transferred to 50 mL polypropylene tubes (SARSTEDT, Newton, NC, USA). Then, the diluted wastewater samples were spiked with 200 µL of 10,000 mg/L of each target compound to reach a final concentration of 50 mg/L. Another set of the control samples were spiked with 200 µL of methanol. The tubes were sealed, shaken, and sit on the bench at ambient temperature for 24 h. After 24 h, RNA was extracted immediately from raw samples, and viral concentrations were quantified by RT-qPCR.

The suppression rates (SR) were calculated using Eq. (2):

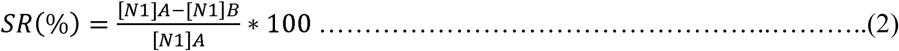

where [N1]_A_ and [N1]_B_ (copies/ µL) are the SARS-CoV-2 concentration in the control (no chemical added) and in the treatment respectively.

#### 2.10.1. Suppression Kinetics

The suppression of SARS-CoV-2 genetic material in wastewater over time was also investigated. The experiments were conducted at room temperature. The spiked wastewater samples (with 50 mg/L of each compound) were collected at times: 0, 3, 6, 12, 24, 48, and 96 hrs. The samples were immediately extracted and processed by RT-qPCR. The dissipation data were fit to the first and second-order kinetic models:

##### First-Order Rate Law

If the rate of reaction exhibits first-order dependence on the concentration of one reactant (*C*), the rate law is expressed as:

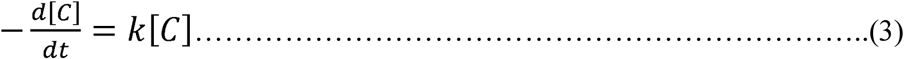

where [*C*] is the concentration of reactant *C*, *k* is the first-order rate constant, and *t* is time. Rearranging the rate law and solving the integral using initial conditions of *t* = 0 and *C* = *C_0_*, the following expression can be found:

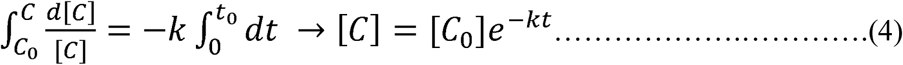

Subsequently, this expression can be written as *ln*[*C*] = -*kt* + *ln*[*C*] Plotting the natural logarithm of the concentration [*C*] versus *t* for a particular reaction will, therefore, allow determination of whether the reaction is first-order. If the reaction is first-order, the slope of the resulting line yields the rate constant *k*. The half-life (*t*_1/2_) of the reaction is given by:

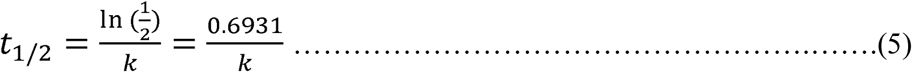

##### Second-Order Rate Law

If the reaction is greater than first-order, the rate law is expressed as:

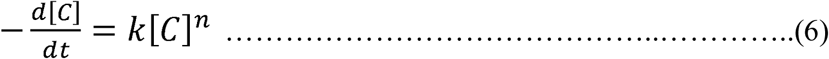

After integrating, the following equation can be obtained:

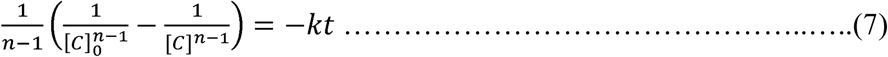

For the second-order reaction (n= 2), both with respect to *C* and overall, the rate law is expressed as:

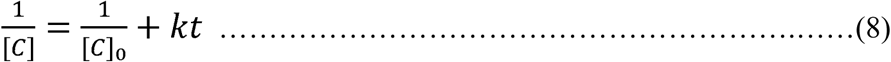

The half-life (*t*_1/2_) of the reaction is given by:

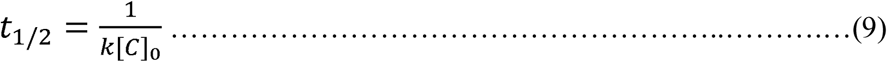

For a second-order reaction involving a reactant, the rate constant *k* can be determined by plotting 1/[C] versus (*t*) to yield a straight line with a slope of *k*.

## 3. Results and Discussion

### 3.1. Identification of the Facilities with High Suppression Rates

Between July 2020 and December 2020, more than fifty-seven wastewater treatment facilities across the state of Missouri, USA were monitored weekly for SARS-CoV-2. This extensive testing of wastewater treatment facilities has provided a comprehensive overview of signal intensity from COVID patients in wastewater. The long-term monitoring showed a clear correlation between the number of COVID patients in a sewershed and the level of viral load in the wastewater (**Figure 1**). However, there is also clear variability among treatment facilities. Specifically, some facilities consistently have lower recovery rates of SARS-CoV-2 load per diagnosed case, suggesting suppression of the genetic material in the sewershed.

**Figure 1.**
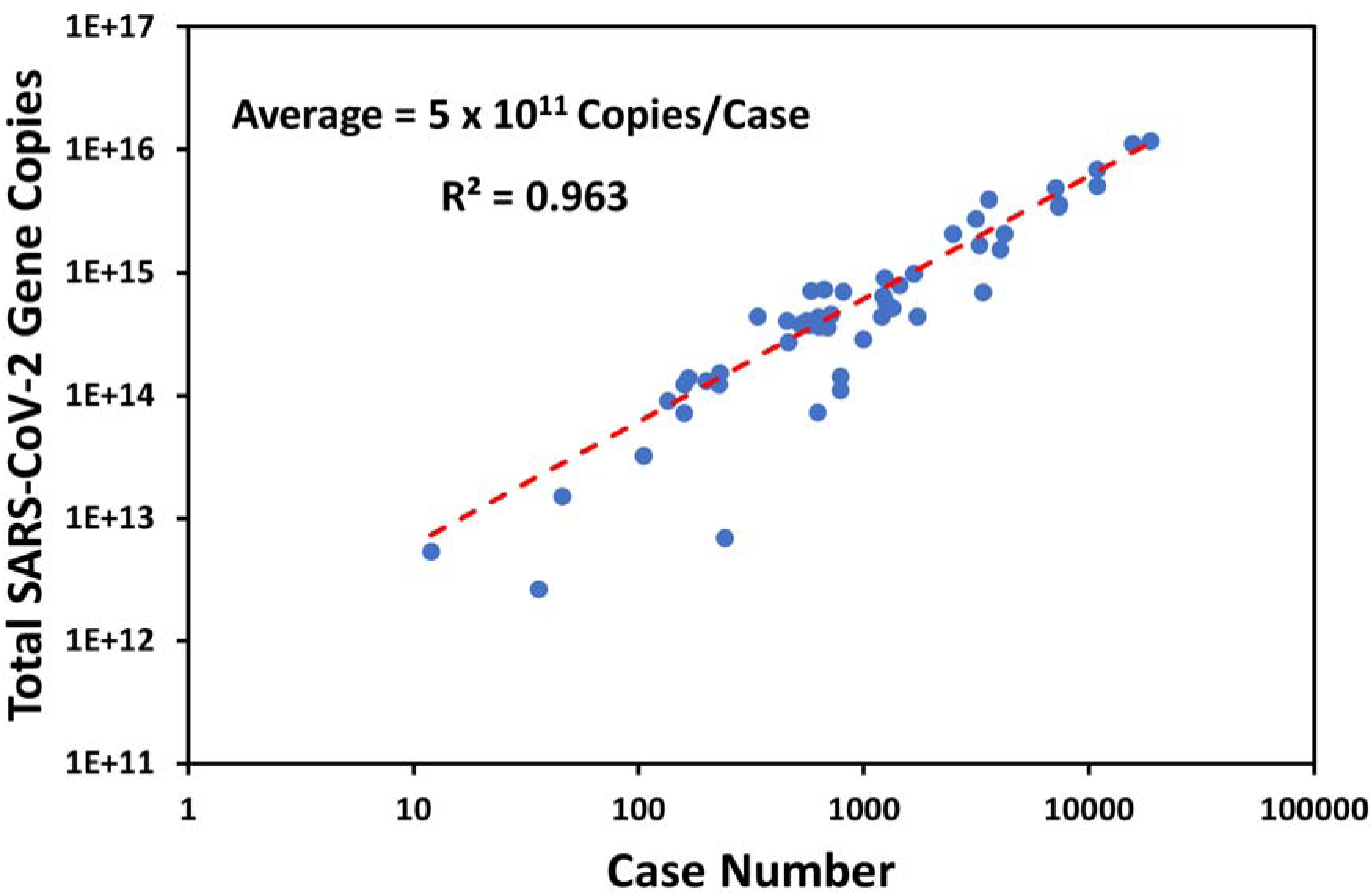
Average SARS-CoV-2 gene copies per diagnosed case. Each data point represents a Missouri wastewater treatment facility (WWTF) from our study. Y-axis is the calculated RNA in the sewershed over the testing period using Eq. (1). X-axis equals the total number of COVID-19 patients identified in each sewershed over the same period.

With data available from MoDNR and DHSS (including reported case numbers), flow rates, along with RT-qPCR results, the average quantity of SARS-CoV-2 load per patient that contributing to the sewershed was calculated (**Figure 1**). The results showed that on average, there are around 5 × 10^11^ SARS-CoV-2 viral load per reported case. Although the amount of SARS-CoV-2 contributed per case varies among communities, there were clear outlier communities that produce little or no genetic material in the wastewater despite the presence of known outbreaks.

**Figure 2** presents the average SARS-CoV-2 viral load per diagnosed case among all the facilities included in this study. According to the results, sewersheds can be divided into three major zones based on SARS-Cov-2 signal suppression (**Figure 2**): Zone 1 includes all the facilities with average viral load/case lower than 5×10^11^ ± 10% variations. These facilities consistently have low recovery rates of viral load per diagnosed case, which suggests viral genetic material suppression in the wastewater. Suppression of viral genetic material in the wastewater could explain the results of Ahmed et al [51], in which no correlation was found between viral genetic material and daily reported cases.

**Figure 2.**
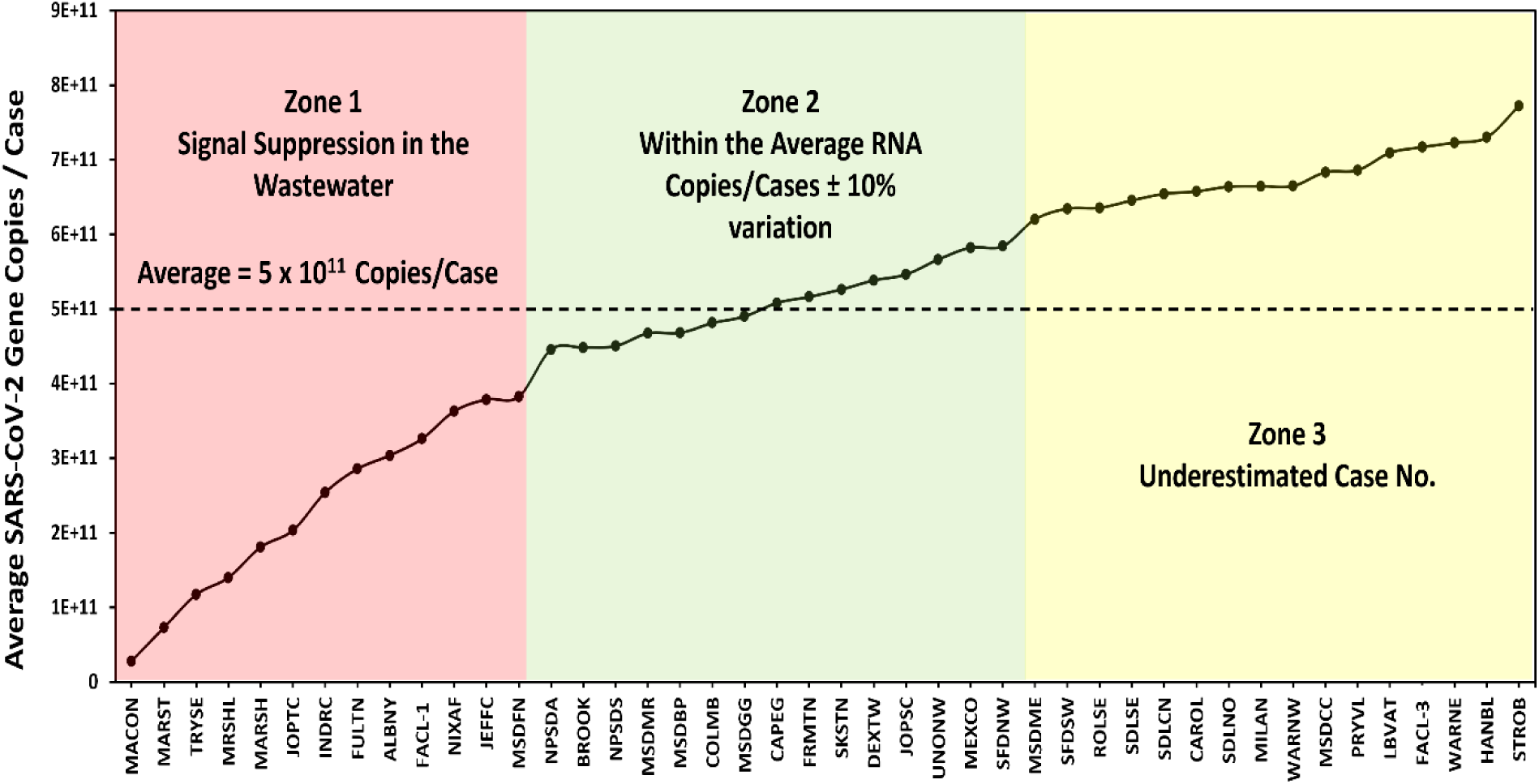
Average SARS-CoV-2 gene copies /case among wastewater treatment facilities (WWTFs). Zone 1 represents facilities with signal suppression; Zone 2 represents the facilities within the average SARS-CoV-2 gene copies/case; Zone 3 represents the facilities with underestimated case number. The abbreviation for each facility are listed in the **Table S1.**

Zone 2 consists of the facilities within the average SARS-CoV-2 load/case (no suppression or signal enhancement). Finally, Zone 3 is comprised of the facilities that have higher numbers of average viral load/case than the predicted values, indicating a likely underestimate in the number of COVID patients. Unreported cases are considered one of the major reasons for average SARS-CoV-2 gene copies being higher than the corresponding case number. During the early phase of the pandemic, clinical testing was limited to multiple criteria, including symptoms and close contacts with a positive case [51]. From these results, among the 57 ranked facilities according to their suppression rates, eight facilities with different suppression rates were chosen for untargeted and targeted analysis (**Figure 3**, and **Table 1**). Six facilities with a range of suppression rates were chosen, including Macon WWTP (MACON), MSD Missouri River WWTP (MSDMR), MSD Fenton WWTP (MSDFN), Independence Rock Creek WWTP (INDRC), Joplin Turkey Creek WWTP (JOPTC), and MSD Bissell Point WWTP (MSDBP). Furthermore, two other facilities with no suppression were included in this study and used as a control: Columbia WWTP (COLUMB) and MSD Grand Glaize WWTP (MSDGG).

**Figure 3.**
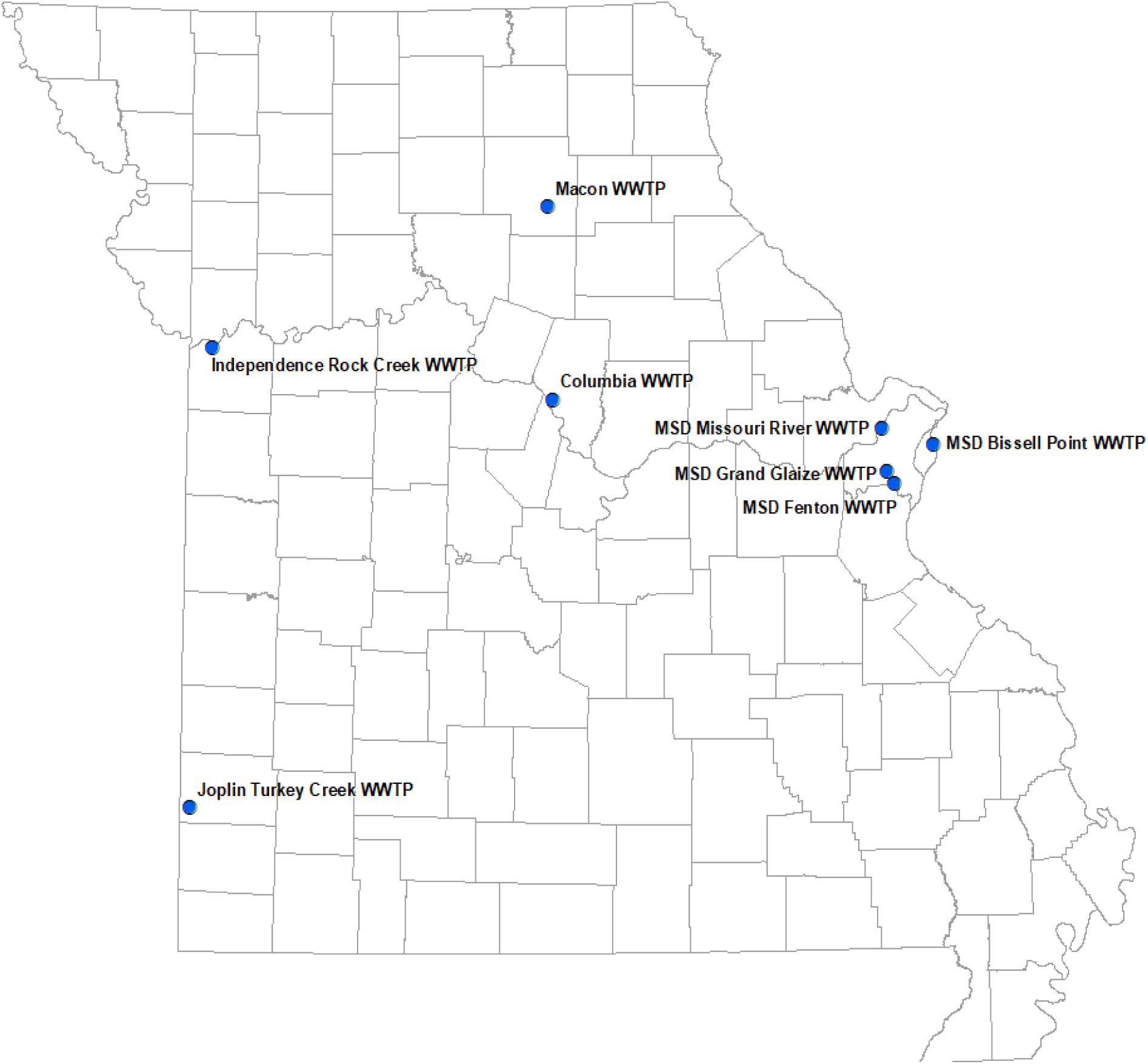
Location of wastewater treatment facilities included in the suppression study.

### 3.2. Untargeted Analyses for Wastewater Extracts

The total ion chromatograms as well as the spectra of active compounds in the wastewater extracts were captured from liquid chromatography-high resolution MS (LC-HRMS) studies. The raw data were processed with the XCMS online platform and the features were annotated using the METLIN library, which resulted in the putative identification of 30 compounds (**Table 2**). These compounds are used for a variety of products such as surfactants, bleaching agents, emulsifiers, and stabilizers (**Table 3**). Heatmap visualization of the clustering of chemical profiles is based on the 30 most significant compounds identified by using a t-test (p < 0.001) (**Figure 4**). Twenty-three compounds exhibited higher relative intensities in suppressed facilities compared to control facilities, contributing significantly to the distinction between control (non-suppression) and suppression facilities (**Figure 4**). Contribution of the variables was determined by examining the variable importance in projection (VIP) score, calculated from the weighted sum of the square for each partial least squares (PLS) loading of each compound [52]. From the top ten compounds identified by VIP, palmitelaidic acid (PAMA), 4-octylphenol (OCPH), N-undecylbenzenesulfonic acid (NUDS), aluminium dodecanoate (ALDO), and 2-dodecylbenzenesulfonic acid (DCBS) were identified as important compounds that significantly contributed to both control and suppression facilities (**Figure 5A**). To further characterize the differences in the relative intensities, partial least squares-discriminant analysis (PLS-DA), a supervised regression technique for classifying groups from multidimensional data, was performed using MetaboAnalyst. PLS-DA analysis with two principal components (PCs) covered 85% of the total variability of the data (**Figure 5B**), indicating significant differences in chemical profiles in control and suppression facilities. The first principal component (PC1) explained 63.9% of the data variability, whereas the second principal component (PC2) accounted for 21.1% of the total variability of the data set.

**Figure 4.**
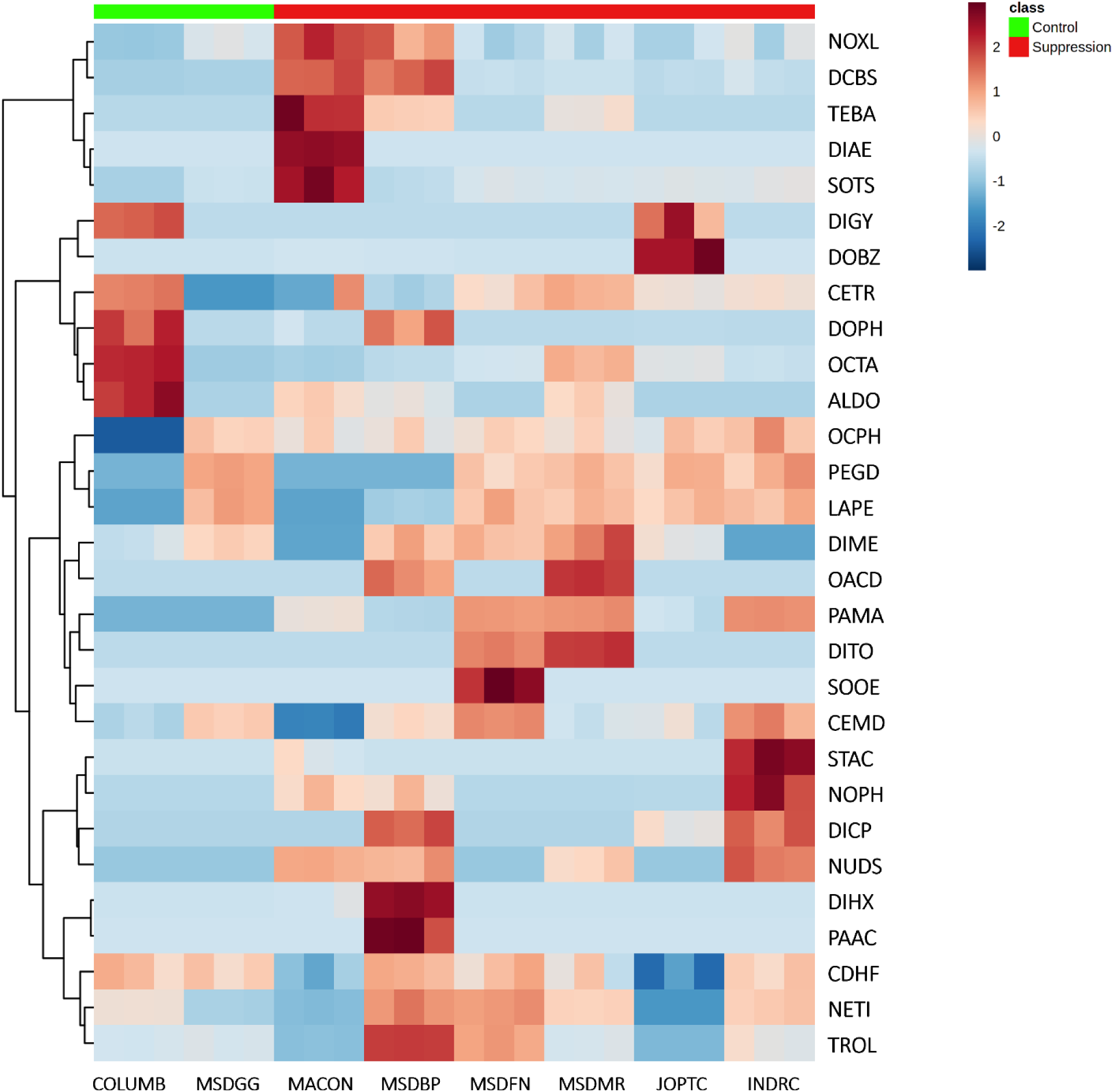
Heatmap of the relative intensities of the identified bioactive found in different locations. Blue represents low relative intensity, whereas red represents high relative intensity. Heatmap features the top thirty metabolite features as identified by t-test analysis (p < 0.001 and intensity ≥10,000). Distance measure is by Euclidean correlation and clustering is determined using the Ward algorithm. The abbreviations of the chemicals are listed in the **Table 2.**

**Figure 5.**
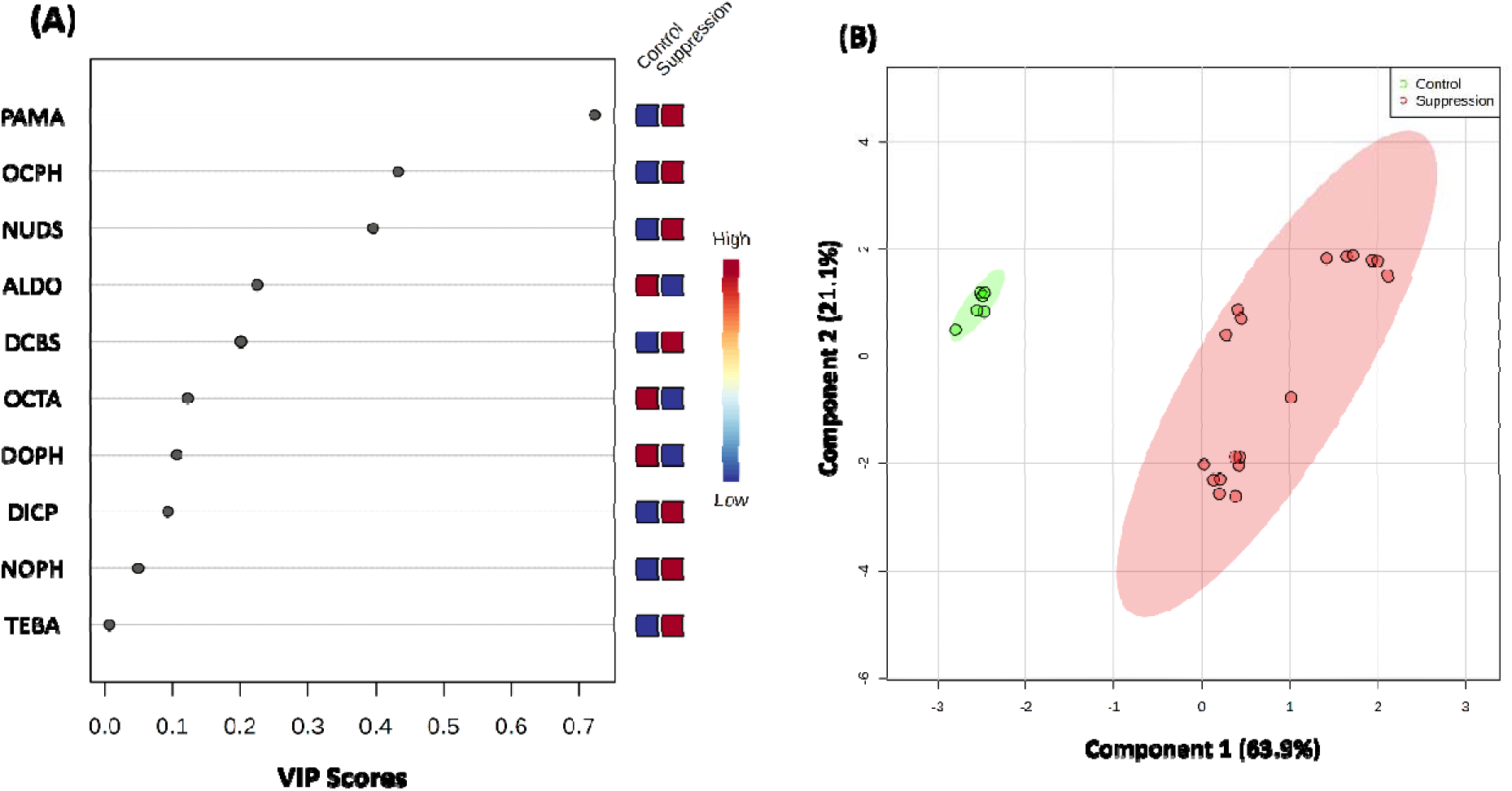
(A) Variable importance in projection (VIP), (B) Partial least squares-discriminant analysis (PLS-DA). In the VIP score plot, the colored boxes indicate the relative intensities of the corresponding compounds in the control and suppression samples. Red represents higher relative abundance, while blue represents lower relative abundance in the VIP score plot. In the PLS-DA plot, the same-colored circles represent replicates of metabolic profiles for each group. The colored ellipses indicate 95% confidence regions of each group.

**Table 2.** List of the compounds putatively identified in the wastewater extracts.

| Putatively identified compound | Abbreviation | Formula | Retention time (min) | Theoretical mass | Extracted mass | Δppm | Adducts |
| --- | --- | --- | --- | --- | --- | --- | --- |
| 4-Octylphenol | OCPH | C<sub>14</sub>H<sub>22</sub>O | 24.21 | 205.1598 | 205.1599 | 0.487 | [M-H] |
| Oleic Acid | OACD | C<sub>18</sub>H<sub>34</sub>O<sub>2</sub> | 32.09 | 281.256 | 281.2494 | 1.422 | [M-H] |
| Lauroyl peroxide | LAPE | C<sub>24</sub>H<sub>46</sub>O<sub>4</sub> | 33.86 | 397.3323 | 397.3325 | 0.503 | [M-H] |
| Palmitic acid | PAAC | C<sub>16</sub>H<sub>32</sub>O<sub>2</sub> | 36.03 | 255.2330 | 255.2331 | 0.392 | [M-H] |
| 2,4-Dichlorotoluene | DITO | C<sub>7</sub>H<sub>6</sub>Cl<sub>2</sub> | 0.52 | 158.9774 | 158.9785 | 6.919 | [M-H] |
| Netilmicin | NETI | C<sub>21</sub>H<sub>41</sub>N<sub>5</sub>O<sub>7</sub> | 4.98 | 476.3079 | 476.3074 | -1.049 | [M+H] |
| Trolamine | TROL | C<sub>6</sub>H<sub>15</sub>NO<sub>3</sub> | 0.56 | 172.0944 | 172.0945 | 0.581 | [M+Na] |
| 3-Chloro-4-(dichloromethyl)-5-hydroxy-2(5H)-furanone | CDHF | C<sub>5</sub>H<sub>3</sub>Cl<sub>3</sub>O<sub>3</sub> | 36.02 | 216.9221 | 216.9229 | 3.687 | [M+H] |
| Dimethicone | DIME | C<sub>6</sub>H<sub>18</sub>OSi<sub>2</sub> | 1.51 | 163.0969 | 163.0967 | -1.226 | [M+H] |
| 4-Dodecylphenol | DOPH | C<sub>18</sub>H<sub>30</sub>O | 27.55 | 280.2635 | 280.2639 | 1.427 | [M+NH<sub>4</sub>] |
| 2-Dodecylbenzenesulfonic acid | DCBS | C<sub>18</sub>H<sub>30</sub>O<sub>3</sub>S | 2.95 | 344.2254 | 344.2261 | 2.033 | [M+NH<sub>4</sub>] |
| Cetrimonium | CETR | C<sub>19</sub>H<sub>42</sub>N | 24.66 | 284.3312 | 284.3318 | 2.11 | [M+H] |
| Diethylene glycol | DIGY | C<sub>4</sub>H<sub>10</sub>O<sub>3</sub> | 32.09 | 107.0703 | 107.0703 | 0 | [M+H] |
| 1-Octadecanamine | OCTA | C<sub>18</sub>H<sub>39</sub>N | 20.39 | 270.3155 | 270.3162 | 2.589 | [M+H] |
| Aluminium dodecanoate | ALDO | C<sub>36</sub>H<sub>69</sub>AlO<sub>6</sub> | 32.2 | 642.5248 | 642.5221 | -4.202 | [M+NH<sub>4</sub>] |
| Dodecylbenzene | DOBZ | C<sub>18</sub>H<sub>30</sub> | 29.88 | 247.2420 | 247.2423 | 1.213 | [M+H] |
| 2-Diethylaminoethanol | DIAE | C<sub>6</sub>H<sub>15</sub>NO | 0.89 | 118.1226 | 118.1225 | -0.846 | [M+H] |
| Palmitelaidic acid | PAMA | C<sub>16</sub>H<sub>30</sub>O<sub>2</sub> | 17.65 | 272.2565 | 272.2567 | 0.734 | [M+NH<sub>4</sub>] |
| Diethanolamine | DEAM | C<sub>4</sub>H<sub>11</sub>NO<sub>2</sub> | 0.59 | 106.0863 | 106.0863 | 0 | [M+H] |
| 4-Nonylphenol | NOPH | C<sub>15</sub>H<sub>24</sub>O | 27.54 | 221.1900 | 221.1908 | 3.617 | [M+H] |
| Polyethylene glycol dioleate | PEGD | C<sub>38</sub>H<sub>70</sub>O<sub>4</sub> | 33.78 | 629.4906 | 629.4928 | 3.494 | [M+K] |
| Dicyclopentadiene | DICP | C<sub>10</sub>H<sub>12</sub> | 27.53 | 133.1012 | 133.101 | -1.502 | [M+H] |
| Nonoxynol-9 | NOXL | C<sub>33</sub>H<sub>60</sub>O<sub>10</sub> | 27.01 | 634.4524 | 634.4551 | 4.255 | [M+NH<sub>4</sub>] |
| Stearic acid | STAC | C<sub>18</sub>H<sub>36</sub>O<sub>2</sub> | 20.19 | 302.3054 | 302.3057 | 0.992 | [M+NH<sub>4</sub>] |
| N-Undecylbenzenesulfonic acid | NUDS | C<sub>17</sub>H<sub>28</sub>O<sub>3</sub>S | 23 | 313.1832 | 313.1838 | 1.915 | [M+H] |
| Dicyclohexylamine | DIHX | C<sub>12</sub>H<sub>23</sub>N | 27.57 | 182.1903 | 182.1902 | -0.548 | [M+H] |
| Tetrabutylammonium | TEBA | C<sub>16</sub>H<sub>36</sub>N | 24.08 | 281.2479 | 281.2482 | 1.066 | [M+K] |
| Sodium Tetradecyl Sulfate | SOTS | C<sub>14</sub>H<sub>29</sub>NaO<sub>4</sub>S | 31.77 | 295.1938 | 295.1948 | 3.387 | [M+H] |
| Sodium oleate | SOOE | C<sub>18</sub>H<sub>33</sub>NaO<sub>2</sub> | 35.01 | 322.2716 | 322.2714 | -0.621 | [M+NH<sub>4</sub>] |
| Cetrimide | CEMD | C<sub>17</sub>H<sub>38</sub>BrN | 3.74 | 358.2080 | 358.2078 | -0.558 | [M+Na] |

**Table 3.** Summary of the compounds screened in the study

| **Compound** | **Usage \*** |
| --- | --- |
| 4-Octylphenol | Soaps, includes personal care products for cleansing the hands or body, and soaps/detergents for cleaning products, homes. |
| Oleic Acid | Surfactants |
| Lauroyl peroxide | Used as bleaching agent |
| Palmitic acid | Wash Aid |
| 2,4-Dichlorotoluene | Antifoaming agents, coagulating agents, dispersion agents, emulsifiers |
| Netilmicin | Aminoglycoside Antibacterial |
| Trolamine | It is found in cosmetics, household detergents, metalworking fluids, polishes and emulsions |
| 3-Chloro-4-(dichloromethyl)-5-hydroxy-2(5H)-furanone | Disinfection byproducts are formed when disinfectants used in water treatment plants react with bromide and/or natural organic matter |
| Dimethicone | Antifoaming |
| 4-Dodecylphenol | Lubricants for engines, brake fluids, oils, refined oil products, fuel oils, etc |
| 2-Dodecylbenzenesulfonic acid | Surfactants |
| Cetrimonium | quaternary ammonium cation whose salts are used as antiseptics. |
| Diethylene glycol | Related to all forms of cleaning/washing, including cleaning products used in the home, laundry detergents, soaps, de-greasers |
| 1-Octadecanamine | Emulsifying; Stabilizing; Surfactant |
| Aluminium dodecanoate | Emulsifier, Stabilizer |
| Dodecylbenzene | Related to all forms of cleaning/washing, including cleaning products used in the home, laundry detergents, soap |
| 2-Diethylaminoethanol | Bleaching agents, cleaning products used in the home, laundry detergents, soaps, de-greasers, spot removers, etc |
| Palmitelaidic acid | Surfactants |
| Diethanolamine | Antistatic; Emulsifying; Foam boosting; Surfactant |
| 4-Nonylphenol | Nonionic detergent metabolite |
| Polyethylene glycol dioleate | Surfactants |
| Dicyclopentadiene | Crude oil, crude petroleum, refined oil products, lubricants for engines, brake fluids, oils |
| Nonoxynol-9 | Surfactants |
| Stearic acid | Surfactants |
| N-Undecylbenzenesulfonic acid | Surfactants |
| Dicyclohexylamine | Related to dishwashing products (soaps, rinsing agents, softeners, etc) |
| Tetrabutylammonium | quaternary ammonium, Household Products, Detergents |
| Sodium Tetradecyl Sulfate | Surfactants |
| Sodium oleate | Surfactants |
| Cetrimide | Preservatives |

### 3.3. Targeted Analyses for Confirmation and Quantification

The molecules tentatively identified through global metabolomic profiling analysis were further confirmed and quantified by LC-MS/MS targeted analyses. Authentic reference standards were used for unambiguous confirmation of compounds and the absolute quantification of the concentrations for each compound identified in the untargeted analysis approach. Due to the limitations of the instrument and limited availability of chemical references standards, fourteen compounds out of thirty were detected and quantified (**Table 4**) and (**Table 5**). **Table 4** summarizes the molecular ions, product ions, retention times, and ionization modes for targeted LC-MSMS analysis of these compounds. The results showed that most of the bioactive compounds had higher concentrations in the wastewater of facilities exhibiting SARS-CoV-2 signal suppression than the control facilities. Four compounds had much higher concentrations in the suppression facilities than the control facilities. In particular, 4-nonylphenol, palmitelaidic acid, sodium oleate, and polyethyleneglycol dioleate exhibited concentrations that were 73.3%, 35.3%, 54%, and 58.8% higher in the suppression facilities than the control facilities, respectively (**Figure 6**). These compounds are mainly used in the production of surfactants and detergents in various industries [53, 54]

**Figure 6.**
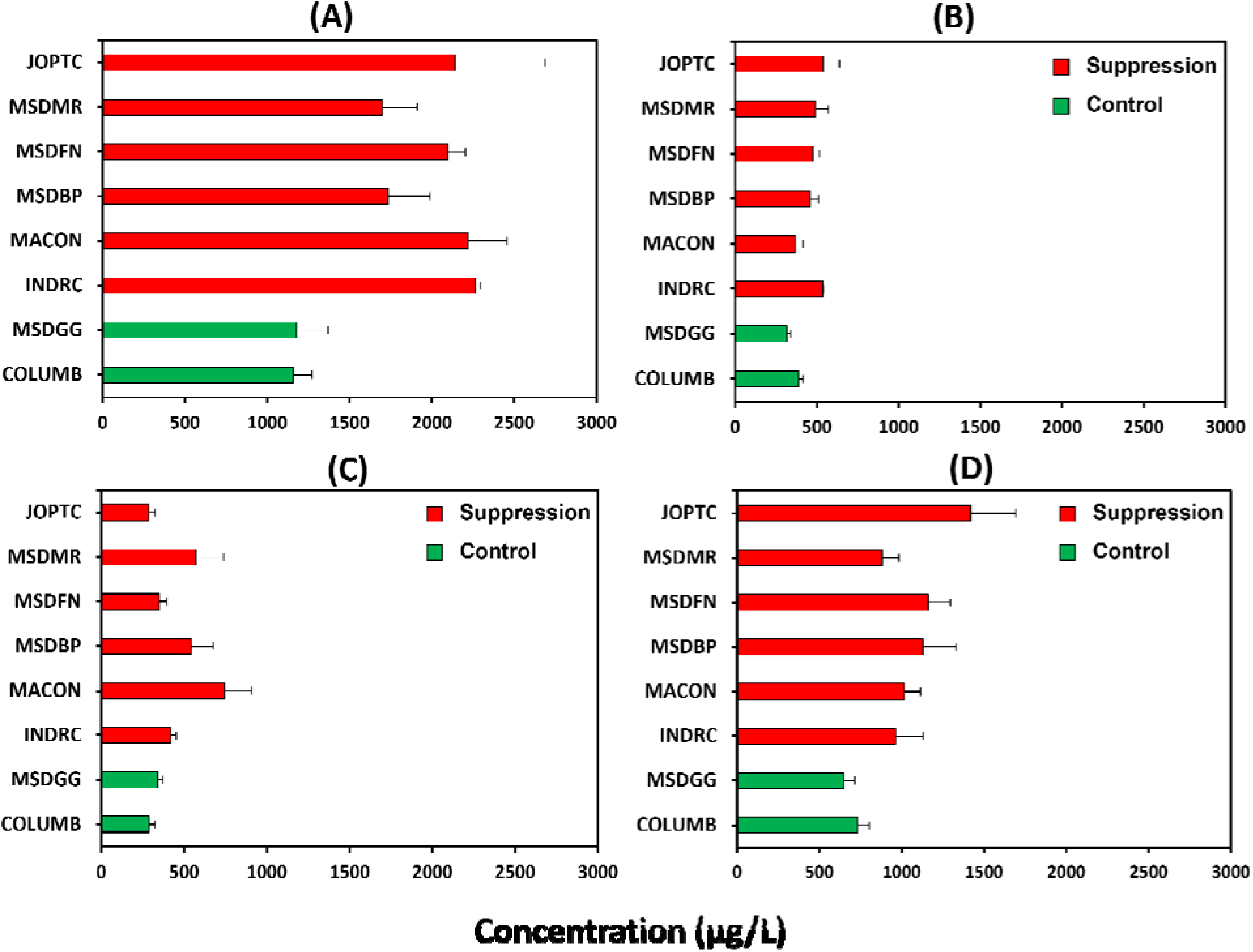
The concentration of the compounds in each facility. (A) 4-nonylphenol; (B) palmitelaidic acid; (C) sodium oleate; (D) polyethylene glycol dioleate

**Table 4.** Molecular and product ions, retention times, and polarity of the compounds identified in wastewater

| Compound | Molecular Ion<br>(m/z) | Product Ion<br>(m/z) | RT (min) | ESI+/ESI- |
| --- | --- | --- | --- | --- |
| 1-Octadecanamine | 242.25 | 56.9 | 12.69 | ESI+ |
| Diethanolamine | 106.1 | 88 | 2.19 | ESI+ |
| 2-Diethylaminoethanol | 118.2 | 72 | 2.13 | ESI+ |
| Dicyclohexylamine | 182.3 | 83 | 7.16 | ESI+ |
| Nonoxynol-9 | 265.3 | 89 | 11.9 | ESI+ |
| Dodecylbenzenesulfonic acid | 325.2 | 183.15 | 10.73 | ESI- |
| Oleic acid | 281.2 | - | 12.11 | ESI- |
| Lauroyl peroxide | 255.3 | 237.5 | 12 | ESI- |
| Palmitic acid | 255.1 | 254.4 | 11.9 | ESI- |
| Stearic acid | 283.3 | - | 12.02 | ESI- |
| 4-Nonylphenol | 219.14 | 133.2 | 12.1 | ESI- |
| Palmitoleic acid | 253.24 | 252.8 | 12.8 | ESI- |
| Sodium oleate | 281.4 | - | 11.72 | ESI- |
| Polyethylene glycol dioleate | 309.3 | 308.6 | 12.29 | ESI+ |
| 4-Octylphenol | 205.16 | 116.55 | 2.9 | ESI- |
| Sodium Tetradecyl Sulfate | 293.06 | 96.85 | 4.36 | ESI- |
| Diethylene glycol | 129.02 | 72 | 2.6 | ESI+ |
| Netilmicin | 476.3 | 190.93 | 0.38 | ESI+ |
| Dicyclopentadiene | 132.1 | - | 0.46 | ESI+ |

**Table 5.** Concentrations of the identified compounds (ppb= µg/L) in influent of each wastewater treatment facility.

| **Compound** | **COLUMB<sup>a</sup>** | **MSDGG<sup>a</sup>** | **INDRC<sup>a</sup>** | **MACON<sup>a</sup>** | **MSDBP<sup>a</sup>** | **MSDFN<sup>a</sup>** | **MSDMR<sup>a</sup>** | **JOPTC<sup>a</sup>** |
| --- | --- | --- | --- | --- | --- | --- | --- | --- |
| 1-Octadecanamine | 143±4.2 | 80.22±10.3 | 346.3±36 | 315±21.1 | 196.82±8.8 | 73.27±5.6 | 71±14.3 | 201.32±24.2 |
| Diethanolamine | 197.19±2.5 | 286±56.7 | 293.36±21 | 145.32±11.4 | 830.32±10.5 | 555.47±21.2 | 201.6±18.8 | 1515±114 |
| 2-Diethylaminoethanol | 43.54±1.8 | 34.78±6.8 | 44.46±5.6 | 570.28±14.2 | 65.16±8.2 | 36.4±1.2 | 35.92±9.2 | 36.48±2.6 |
| Dicyclohexylamine | 0.63±0.05 | 1.1±0.1 | 0.72±0.04 | 1±0.09 | 67.55±3.8 | 0.85±0.1 | 0.7±0.08 | 0.82±0.2 |
| Nonoxynol-9 | 470.1±74.1 | 353.6±69.1 | 356.2±10.5 | 1120.7±22.1 | 548.6±65.2 | 617.16±22.8 | 532.31±15.8 | 323.77±21.1 |
| Dodecylbenzenesulfonic acid | 1510.46±140.8 | 1647.18±93.2 | 1459.32±60.2 | 578±25.1 | 907.45±66.1 | 1563.43±33.1 | 1833.8±22.2 | 1198.93±20.1 |
| Oleic acid | 939.34±56.8 | 495.62±47.2 | 421.51±12.5 | 712.94±32.1 | 360.25±23.4 | 221.52±14.1 | 560.71±100.2 | 422.54±21.4 |
| Lauroyl peroxide | 4860.82±215 | 4333±151 | 5106±105 | 11017±111 | 7759.6±212 | 2176.6±20.4 | 2391.2±25.4 | 5383.6±23.5 |
| Palmitic acid | 3279±165 | 2142±64.2 | 3886.2±110 | 3769.2±214 | 3114.1±215 | 1667.8±65.2 | 1840±36.5 | 4041.3±132.2 |
| Stearic acid | 1621.7±89.4 | 1692.6±85.2 | 1751±36.5 | 1649.3±125 | 1541.6±111.2 | 1346.5±135.1 | 1536.8±121 | 1652.1±32.5 |
| 4-Nonylphenol | 1159.6±110.5 | 1432.5±190.2 | 2263.31±31.8 | 2220.28±233.8 | 1733±255.2 | 2095.5±107.6 | 1699.8±211.7 | 2142.4±543.4 |
| Palmitelaidic acid | 570.3±51.7 | 317.3±21.7 | 535.43±5.6 | 369.7±44.6 | 458±51.7 | 476±40.5 | 491.7±78.5 | 537.75±98.8 |
| Sodium oleate | 288.15±35 | 340.33±32 | 417.71±35.5 | 742.3±163.5 | 543.6±131.2 | 349.5±46.5 | 570.1±167.1 | 286.1±38 |
| Polyethylene glycol dioleate | 1182.2±284.6 | 873.8±138 | 963.1±169 | 1014.86±99.7 | 1128.32±202.2 | 1162.42±133.5 | 883.64±99.7 | 1418.75±274 |

<sup>a</sup> Absolute concentrations were determined by LC-MS/MS with authentic standards.

The concentrations of 4-nonylphenol in the urban wastewaters were determined in Japan, China, and USA. The concentrations were about 190 µg/L [55], 2 µg/L [56], and 400 µg/L [57], respectively. In this study, average concentrations of 4-nonylphenol were 1169 ± 13.3 µg/L and 2025.7 ± 247 µg/L in the control and suppression facilities, respectively. No information was found regarding the concentrations of the other three compounds in wastewater. Palmitelaidic acid was reported to be used to produce cosmetics, soaps, and industrial mold release agents [58]. and the average concentrations were 353.4 ± 51.2 µg/L and 478.2 ± 62 µg/L in the control and suppression facilities, respectively. According to the Consumer Product Information Database (CPID), polyethylene glycol dioleate (PEDG) is used as surface active agent and lubricant additive in different kinds of household and commercial products (e.g., stainless steel cleaner & polish, wood polish)[59]. The average concentration of PEGD in the control facilities was 689.3 ± 58.4 µg/L, while the average concentration in the suppression facilities was 1095.2 ±189.2 µg/L. Finally, sodium oleate is one of the major ingredients of metal polishes and is also used as an emulsifier in the polymerization of different compounds, according to Hazardous Substances Data Bank (HSDB)[60]. The observed concentrations of sodium oleate were 314.2 ± 37 µg/L and 485 ± 183 µg/L in the control and suppression facilities, respectively.

The presence of different industries in the sewersheds served by the suppression facilities might be the reason behind the high concentrations of these surfactants in the wastewater (**Table 6**). For example, the majority of the sewersheds contain food processing, cleaning products, plastics, and fabrics, and metal finishing industries which can significantly contribute chemicals to the wastewater received by the investigated facilities. Several studies have been done on the monitoring of wastewater for different compounds used as surfactants and detergents [31,37,61– 64]. However, there was no study on the effect of these compounds on SARS-CoV-2 in the wastewater. Thus, in the next section, the stability of SARS-CoV-2 genetic material in wastewater in the presence of four compounds is discussed.

**Table 6.** Industrial category found in the sewershed served by suppression facilities.

| **No** | **Facility Name** | **Facility ID** | **Industrial Category** |
| --- | --- | --- | --- |
| 1 | Joplin Turkey Creek WWTP | JOPTC | Food processing and packaging, wood preservation, metal finishing, roofing and building products, hospital |
| 2 | Independence Rock Creek WWTP | INDRC | Metal finishing, food packaging, plastics, car part, Bulk Fuel |
| 3 | Macon WWTP | MACON | Food manufacture, tool, and Dye |
| 4 | MSD Bissell Point WWTP | MSDBP | Fabric, chrome plating, metals finishing, electronics, cleaning products, detergent, leather, food packaging, paper, plastics, polishes, hospital |
| 5 | MSD Fenton WWTP | MSDFN | Packaging, cleaning products |
| 6 | MSD Missouri River WWTP | MSDMR | Metals finishing, plastics, paper, electronics, hospital |

### 3.4. Identification of the Bioactive Molecules Associated with Suppression of SARS-CoV-2 Signals

To further characterize the findings from the metabolomic approach, stepwise regression models and LASSO regression models were used to determine the significant predictor variables (i.e., compounds’ relative intensities) which are positively correlated with the response variable (i.e., SARS-CoV-2 suppression rate). Results from positive and negative ion modes were analyzed separately.

The relationships among chemical signal intensities (generated from UPLC-MS positive ion mode analysis) and SARS-CoV-2 RNA suppression rate were examined using four different statistical approaches. According to the forward and backward stepwise regression models, the signal intensities of 13 out of 21 compounds were positively correlated with the viral suppression rate (**Table S2**). Best subsets regression also identified the signal intensities of 13 out of 21 compounds as being positively correlated with the viral suppression rate (**Table S3**). The signal intensities of eight out of 21 compounds were kept in the lasso regression model and obtained positive estimated coefficients (**Table S4**). Palmitelaidic acid, 4-nonylphenol, dicyclopentadiene, tetrabutylammonium and sodium oleate signal intensities were positively correlated with the viral suppression rate among all four statistical approaches (**Table S2-S4**). Furthermore, using the same statistical approaches, polyoxyethylene glycol dioleate and 4-nonylphenol appeared to be positive correlated to vial suppression rate among all four approaches when the signal intensities from negative ion mode were analyzed (**Table S5 and S6**). In conclusion, only the signal intensity of 4-nonylphenol was positively correlated with the viral suppression rate for both positive and negative ion modes.

### 3.5. Suppression Experiments

The results from the statistical approaches suggested that the signal intensities of 4-nonylphenol, palmitelaidic acid, sodium oleate, and polyethylene glycol dioleate are positively correlated with SARS-CoV-2 suppression rates (**Table S2-S6**). Therefore, the suppression of these compounds on SARS-CoV-2 were tested in incubation studies using real wastewater. A wastewater with known high viral copy numbers from “non-suppressed” facilities was used in these experiments. **Figure 7** shows the suppression rates (SR) of the compounds tested. After reacting for 24h, the SR (%) were 57.2%, 35%, 43.3%, and 78.2% when adding PEGD, NOPH, SOOE, and PAMA, respectively.

**Figure 7.**
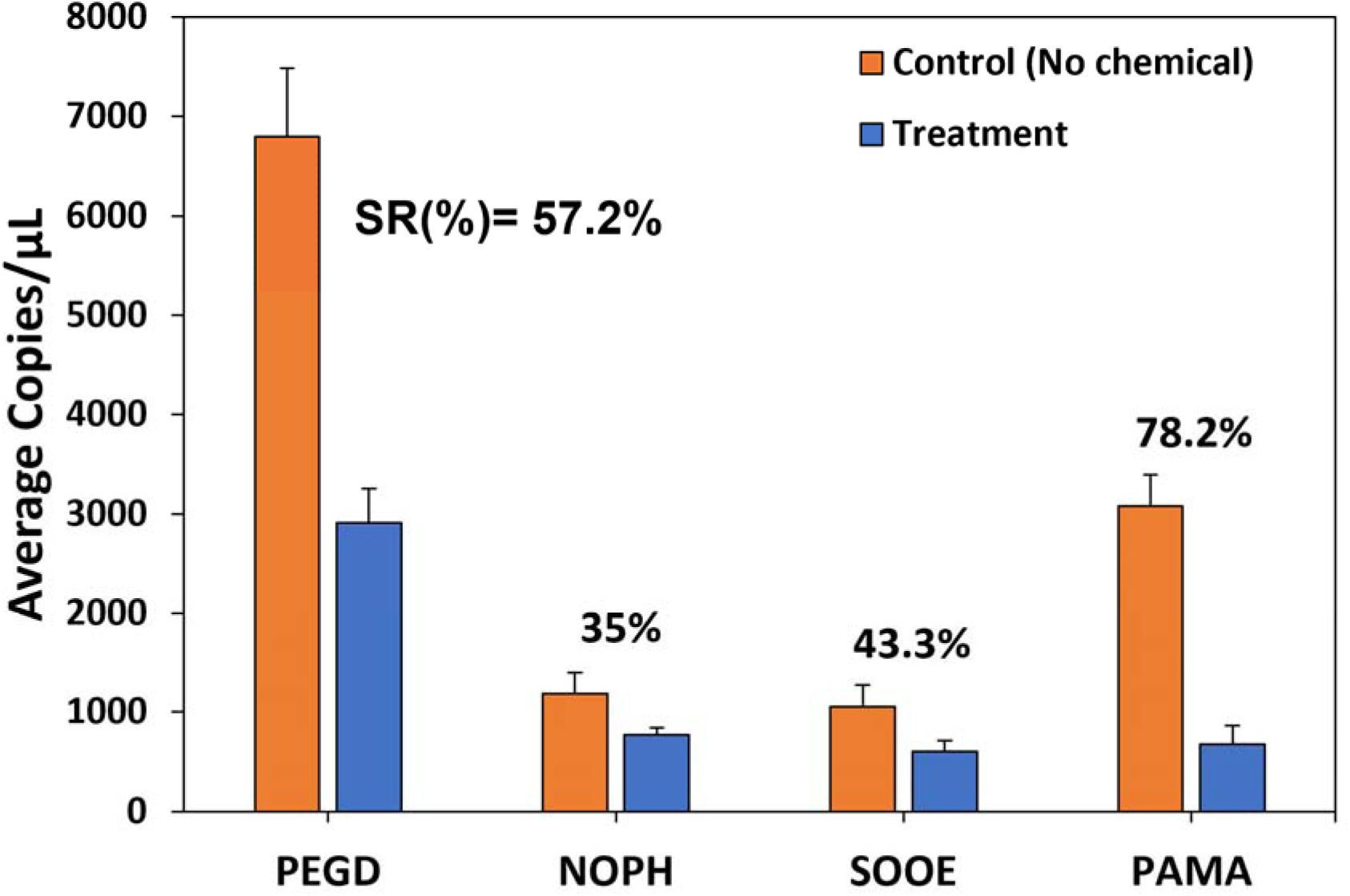
Chemical effect on the SARS-CoV-2 signals in the wastewater. Samples from different batches were treated with 50 mg/L PEGD (polyethylene glycol dioleate), NOPH (4-nonylphenol), SOOE (sodium oleate), and PAMA (palmitelaidic acid). Wastewater samples were reacted with each chemical individually for 24 h at room temperature. Error bars represent standard deviation.

Enveloped viruses like SARS-CoV-2 have a variety of sites on the lipid membrane/envelop embedded with proteins where surfactants (nonionic, anionic, and cationic surfactants) can bind and interact [65]. In general, surfactants are well known to bind to proteins, with the main mechanisms being hydrophobic, electrostatic, and H-bonding. The binding of the surfactants often leads to denaturation of the protein, either by the formation of protein-surfactant complexes or by unfolding [65, 66].

For enveloped viruses, a major point of attraction to surfactant molecules is the lipid bilayer in which hydrophobic interaction may become the main driving force. In addition to hydrophobic interactions, electrostatics may also play a role, especially if the surfactant was oppositely charged [65]. Some surfactants might be bound within the lipid bilayer and this binding will raise the chemical potential of the surfactant in the bilayer, leading to thermodynamic instability[67]. The four compounds tested were considered hydrophobic because their partitioning coefficient (logP) ranges between 5.6-15, demonstrating that hydrophobic interaction plays an important role in the interaction between surfactants and lipid bilayers.

The suppression of SARS-CoV-2 RNA in wastewater over time was also investigated. The experiments were conducted at room temperature. Spiked wastewaters (with 50 mg/L of each compound) were collected at the following times: 0, 3, 6, 12, 24, 48, and 96 hrs. Samples were immediately extracted and processed by RT-qPCR. **Figure 8** shows the kinetic experimental results for both palmitelaidic acid (PAMA) and polyethylene glycol dioleate (PEGD). For both figures (A and B), the data are normalized by the number of RNA copies/µL in the control samples at time zero. PAMA and PEGD suppressed 70% and 65% of SARS-CoV-2 RNA for the first 6 hrs of the experiment, respectively. From our observation, the two compounds immediately suppressed the genetic material in the wastewater, and as such, the existence of these two compounds at 50 mg/L will dramatically decrease the COVID-19 signals in wastewater. It is therefore critical to determine the real concentrations of the compounds that reduce the stability of the genetic material signals in wastewater. Based on the known concentrations, correction factors may be developed to achieve more reliable and unbiased surveillance results for wastewater treatment facilities receiving wastewater from industries.

**Figure 8.**
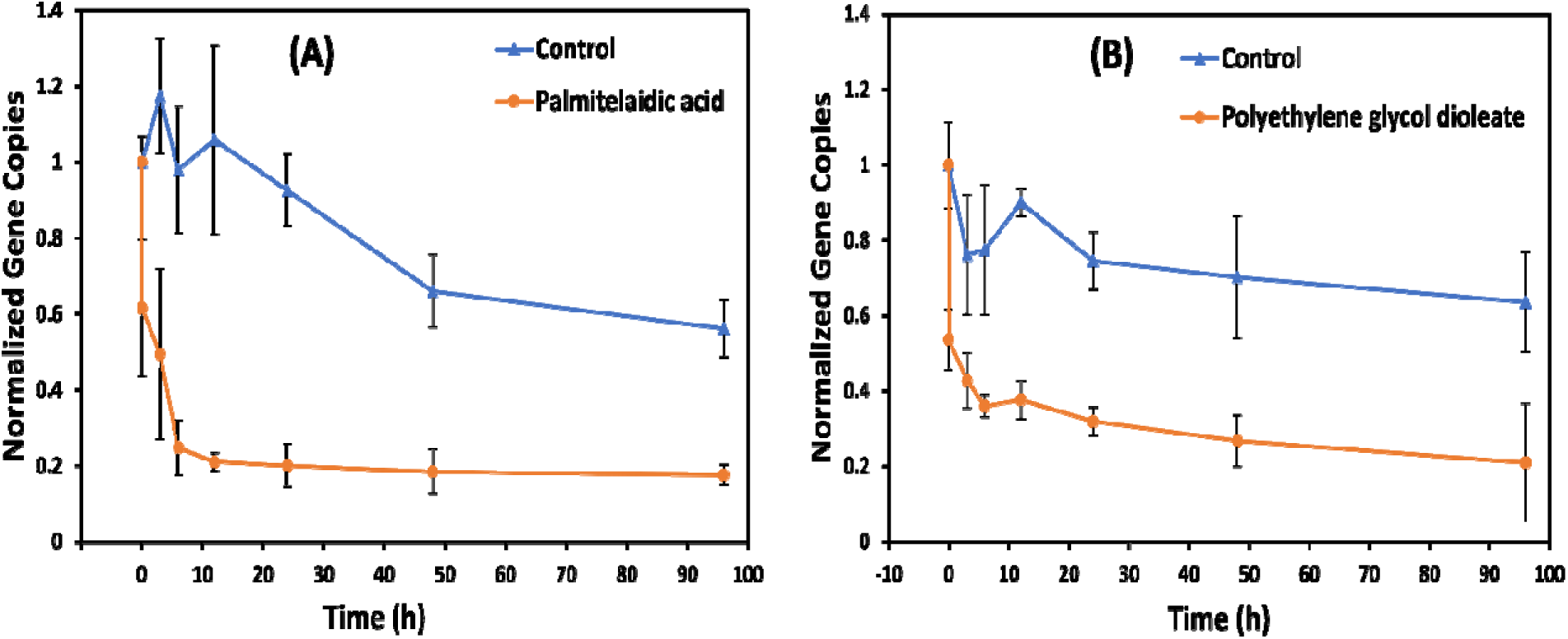
Kinetic experiments. (A) 50 mg/L of palmitelaidic acid spiked into wastewater with known SARS-CoV-2 gene copies. (B) 50 mg/L of polyethylene glycol dioleate spiked into wastewater with known SARS-CoV-2 gene copies. The wastewaters were from two different batches. All the experiments were conducted at room temperature. The collected samples were immediately extracted and processed by RT-qPCR.

In order to calculate the rate constant of the reaction (*k*) and the half-life of the viral RNA (*t_1/2_*), the data from **Figure 8** was used to determine the order of the reaction. Zero-order, first-order, and second-order were tested and the results showed that all the data fit the second-order reaction (**Figure 9**). This meant that the rate of the reaction increases by the square of the increased concentration of the SARS-CoV-2 RNA in the wastewater. The calculated half-lives were compared to the results from 24 h (**Figure 7**). The SARS-CoV-2 RNA were suppressed by 78.2% and 57.2% when adding PEGD and PAMA, respectively. The calculated t_1/2_ (the time when SARS-CoV-2 concentrations drop to its half value) were 8.5 h and 2.2 h for PEGD and PAMA respectively (**Figure 9**). In an effort to evaluate the role that well-shaking plays in the half-life calculation, another experiment was conducted on the rocker at room temperature. Samples were continuously agitated during the experiment period. The constant agitation on the rocker (Fisherbrand^TM^ Nutating Mixers, PA, USA) at 10 rpm was supposed to provide a homogenous mixture and allow the compounds to interact with SARS-CoV-2. The data from mixing experiments were also fit the second-order reaction (**Figure 11**). The 24 h results showed that the suppression rates for SARS-CoV-2 RNA were 39.2% and 45% and the calculated t_1/2_ were 16.4 h and 10.3 h, when PEGD and PAMA were added, respectively. Surprisingly, the calculated t_1/2_ in the mixing experiments for both compounds were higher than the sitting condition. During the agitation, the suspended solids in the wastewater were competing for the reactive chemicals, and as a result, a lower concentration is available to interact with SARS-CoV-2, leading to longer times to reach *t_1/2_*.

**Figure 9.**
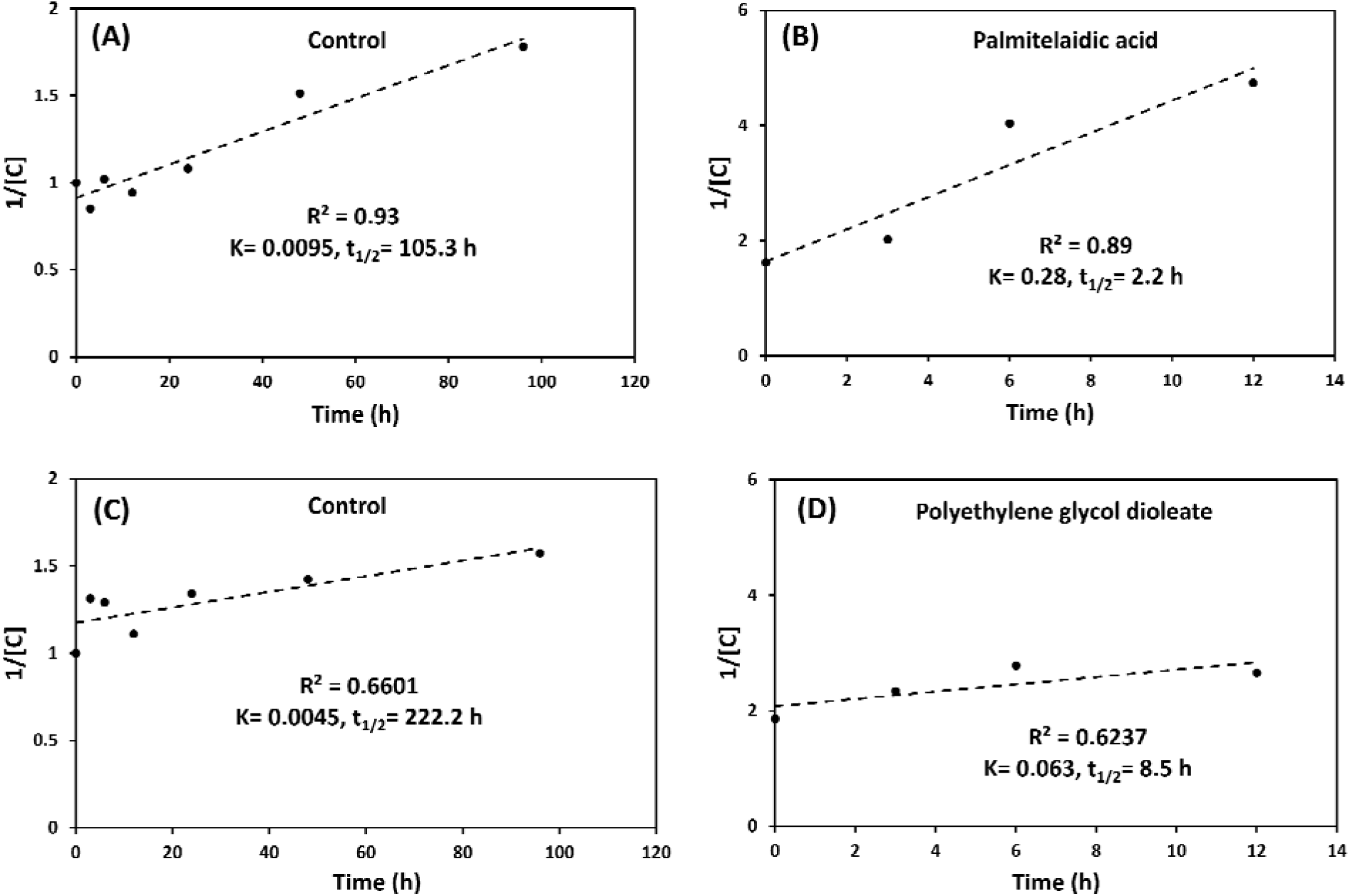
Rate constant and half-life of the reaction. (A) Control for PAMA. (B) Palmitelaidic acid (PAMA). (C) Control for PEGD. (D) Polyethylene glycol dioleate (PEGD). The wastewaters were from two different batches. All the experiments were conducted at room temperature. All samples were sitting still on the bench and no agitating was involved.

**Figure 10.**
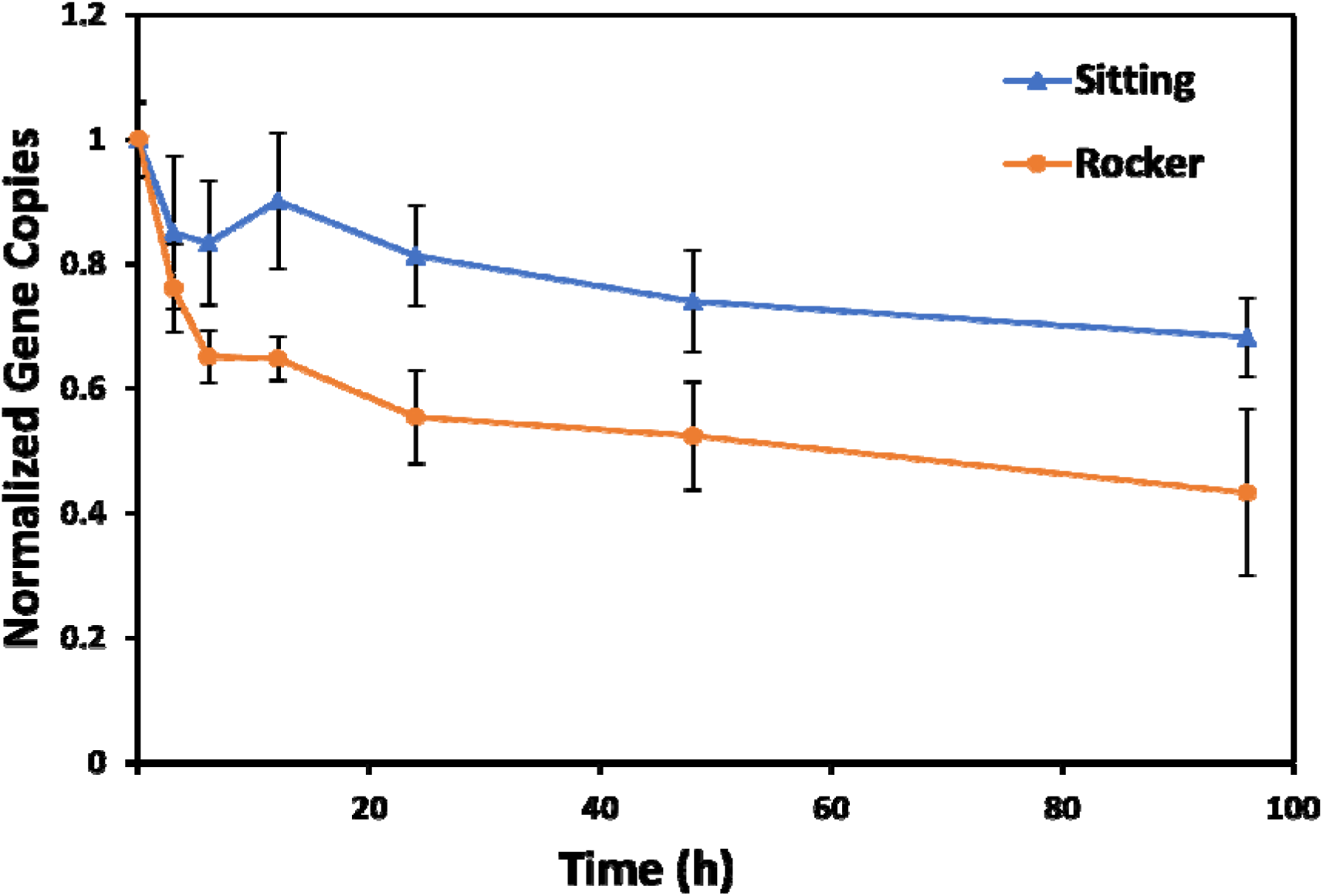
Kinetic experiments for control samples. No chemicals were spiked. The wastewaters were from two different batches. The data were normalized by the gene copies/µL at time zero. All the experiments were conducted at room temperature. The collected samples were immediately extracted and processed by RT-qPCR.

**Figure 11.**
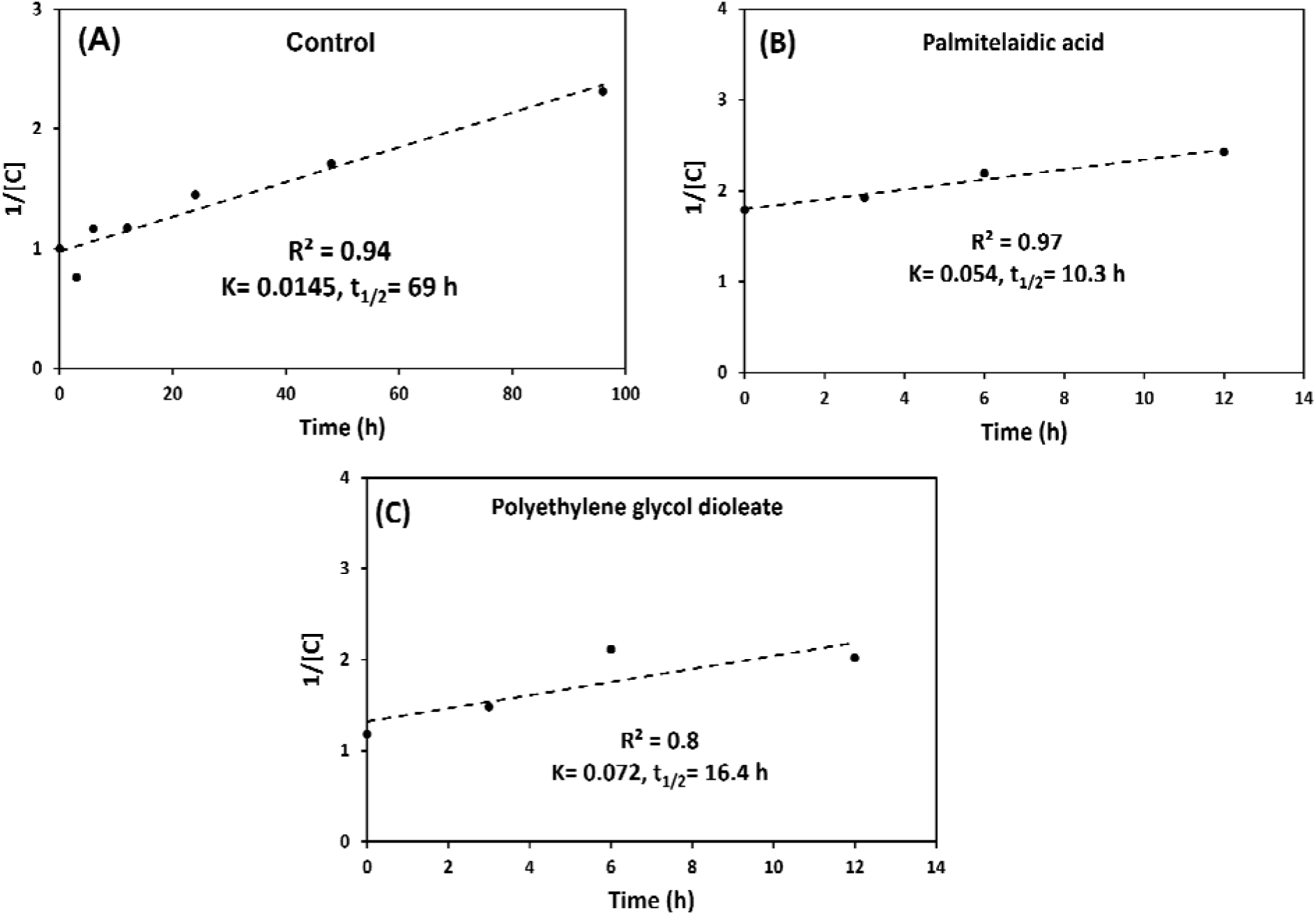
Rate constant and half-life of the reaction. (A) Control. (B) Palmitelaidic acid (PAMA). (C) Polyethylene glycol dioleate (PEGD). The wastewater waw from one batch. All the experiments were conducted at room temperature. All the samples were continuously agitated on the rocker.

It is also important to mention that SARS-CoV-2 in the control samples was less stable when agitating over time compared to sitting samples (**Figure 10**). The half-life of the viral RNA reflected this observation, the average *t_1/2_* for the control samples with no agitating was 163 h and was about 69 h when the control samples were continuously agitated (**Figure 11**). The main reason that could explain this observation is that the agitating process will allow the chemicals present in wastewater to interact with SARS-CoV-2, resulting in a shorter half-life. Furthermore, this finding might explain the conflicted findings reported among different studies. For example, Robinson et al [68](Missouri team), the Ohio State [69], and the team at University of Notre Dame [70] reported constant stability of SARS-CoV-2 in wastewater at room temperature for at least 5-7 days, while the findings reported by Weidhaas et al [27] (team from Utah) suggest rapid degradation of the SARS-CoV-2 signal following a first order decay constant at both 4 C, 10 C, or 35 C within 24 h, with the virus signal not being detectable after 12 h of storage at 35 °C. Similar susceptibility to decay and degradation of SARS-CoV-2 RNA by increasing temperature in wastewater were also reported by Ahmed et al. [71]. The accelerated transfer of energy resulting from the mixing process has demonstrated the similar temperature effects on the stability of SARS-CoV-2 signals.

## 4. Conclusions

Approximately 20% of our currently tested wastewater treatment facilities (WWTFs) in Missouri, USA receive some input from industries. Several classes of molecules released by these regional industries and manufacturing facilities, particularly the food processing industry, significantly suppressed the signals of SARS-CoV-2 in wastewater by breaking down the lipid-bilayer of viral membranes. By taking advantage of recent advancements in mass spectrometry, metabolomics algorithms, computational capacity and mass spectral reference databases, we have successfully identified and quantified several bioactive chemicals that suppress the signals of the SARS-CoV-2 in wastewater. The chemical suppressors include active ingredients in surfactants, detergents, lubricants, preservatives, degreasers, and disinfection products. Based on the concentrations of these bioactive molecules that significantly reduce the stability of the SARS-CoV-2 genetic markers signals in wastewater (e.g., 4-nonylphenol, palmitelaidic acid, sodium oleate, and polyethylene glycol dioleate), correction factors could be developed to achieve more reliable and unbiased surveillance results for wastewater treatment facilities receiving wastewater from industries. In addition, our findings from the suppression kinetics experiments suggest that the stability of SARS-CoV-2 in wastewater is also strongly influenced by the sample preparation process (i.e., agitating vs. sitting still), which might account for the conflicting findings reported among different studies.

## Data Availability

All data produced in the present study are available upon reasonable request to the authors

## Acknowledgements

The authors would like to thank the Missouri Department of Health and Senior Services (DHSS) administrating the funding. We would like to express our gratitude to the Missouri Department of Natural Resources (DNR) for coordinating the sample collection and to the municipalities and wastewater operators that donated their time to collect samples analyzed in this research. Research reported in this publication was supported by funding from the Centers for Disease Control and the National Institute on Drug Abuse of the National Institutes of Health under award number U01DA053893-01. We would also like to thank the Center for Agroforestry at University of Missouri, USDA/ARS Dale Bumpers Small Farm Research Center under agreement number 58-6020-6-001 from the USDA Agricultural Research Service for supporting part of this research. The content is solely the responsibility of the authors and does not necessarily represent the official views of the National Institutes of Health, the Centers for Disease Control or USDA-ARS.

