## Supplementary Information for "Identification and Quantification of Bioactive Compounds Suppressing SARS-CoV-2 Signals in Wastewater-based Epidemiology Surveillance"

**Table S1.** List of the wastewater treatment facilities in Missouri

| **No.** | **Project ID** | **City** | **Population Served** | ^a^**Facility Capacity** | **Composite Sampling Mode** | **^b^ Average Influent Flow** |
| --- | --- | --- | --- | --- | --- | --- |
| 1 | CARTH | Carthage | 12,000 | 7 | Time Based | 3.82 |
| 2 | ROLSE | Rolla | 15,000 | 4.8 | Time Based | 4.39 |
| 3 | SDLCN | Sedalia | 7,500 | 3.03 | Time Based | 3.03 |
| 4 | KCBLU | Kansas City | 287,500 | 105 | Time Based | 101.47 |
| 5 | SFDSW | Springfield | 173,676 | 64 | Time Based | 48.56 |
| 6 | CASVL | Cassville | 3,300 | 1.1 | Time Based | 1.01 |
| 7 | FACL-1 | - | 12,000 | 9 | Time Based | 2.80 |
| 8 | MONET | Monett | 9,100 | 6 | Time Based | 4.40 |
| 9 | JOPTC | Joplin | 35,462 | 15 | Time Based | 16.10 |
| 10 | WARNE | Warrensberg | 7,990 | 1.5 | Flow Based | 2.29 |
| 11 | CHARL | Charleston | 4,000 | 1.5 | Time Based | 1.50 |
| 12 | ALBNY | Albany | 1,710 | 0.49 | Time Based | 0.19 |
| 13 | MSDME | St. Louis | 451,367 | 210 | Time Based | 97.40 |
| 14 | UNONW | Union | 7,378 | 1.5 | Time Based | 0.75 |
| 15 | FULTN | Fulton | 12,790 | 2.9 | Time Based | 1.90 |
| 16 | MSDGG | Valley Park | 115,895 | 21 | Time Based | 13.61 |
| 17 | FACL-2 | - | 6,155 | 3.4 | Time Based | 1.59 |
| 18 | SFDNW | Springfield | 26,078 | 6.8 | Time Based | 13.60 |
| 19 | MILAN | Milan | 1,960 | 0.7 | Flow Based | 0.38 |
| 20 | HANBL | Hannibal | 16,000 | 12 | Time Based | 4.14 |
| 21 | STPSC | St. Peters | 48,774 | 9.5 | Time Based | 6.51 |
| 22 | JEFFC | Jefferson City | 75,000 | 11 | Flow Based | 8.10 |
| 23 | NPSDS | FENTON | 44,035 | 4 | Time Based | 2.27 |
| 24 | JOPSC | Joplin | 11,800 | 7.2 | Time Based | 6.30 |
| 25 | SDLSE | Sedalia | 7,500 | 2.6 | Time Based | 4.29 |
| 26 | FRMTN | Farmington | 9,525 | 2.75 | Time Based | 2.87 |
| 27 | WARNW | Warrensburg | 6,235 | 1.5 | Flow Based | 2.20 |
| 28 | SDLNO | Sedalia | 8,250 | 2.5 | Time Based | 1.47 |
| 29 | CAROL | Carrollton | 3,784 | 1.5 | Time Based | 0.73 |
| 30 | MACON | Macon | 5,471 | 2.5 | Time Based | 1.12 |
| 31 | SKSTN | Sikeston | 17,000 | 5 | Time Based | 2.40 |
| 32 | MSDBP | St. Louis | 306,647 | 150 | Time Based | 93.80 |
| 33 | STROB | St. Roberts | 4,085 | 1 | Time Based | 0.64 |
| 34 | PRYVL | Perryville | 9,000 | 1.8 | Time Based | 1.09 |
| 35 | DEXTW | Dexter | 2,400 | 0.78 | Time Based | 0.75 |
| 36 | FACL-3 | - | 900 | 0.14 | Time Based | 0.06 |
| 37 | COLMB | Columbia | 123,180 | 20.6 | Time Based | 15.70 |
| 38 | MSDLM | St. Louis | 66,738 | 15 | Time Based | 9.55 |
| 39 | MSDFN | St. Louis | 24,174 | 6.75 | Time Based | 4.05 |
| 40 | MRSHL | Marshall | 13,000 | 7.1 | Time Based | 2.38 |
| 41 | STJOE | St. Joseph | 58,200 | 27 | Time Based | 17.73 |
| 42 | MSDMR | St. Louis | 174,537 | 38 | Time Based | 17.70 |
| 43 | LBVAT | Independence | 360,000 | 52 | Time Based | 85.66 |
| 44 | FACL-4 | - | 10,559 | 5.3 | Time Based | 2.73 |
| 45 | BROOK | Brookfield | 4,600 | 2 | Time Based | 1.12 |
| 46 | CAPEG | Cape Girardeau | 38,000 | 11 | Flow Based | 7.64 |
| 47 | MEXCO | Mexico | 11,500 | 3 | Time Based | 3.40 |
| 48 | MARSH | Marshfield | 8,000 | 1.5 | Time Based | 2.25 |
| 49 | NIXAF | Nixa | 20,000 | 4 | Time Based | 2.00 |
| 50 | NEVAD | Nevada | 3,674 | 2 | Time Based | 2.75 |
| 51 | WARSW | Warsaw | 2,204 | 0.45 | Flow Based | 0.70 |
| 52 | BLIVR | Bolivar | 10,500 | 2.55 | Time Based | 1.37 |
| 53 | ELDON | Eldon | 4,895 | 1 | Time Based | 0.98 |
| 54 | WPLAN | West Plains | 12,000 | 3 | Time Based | 5.42 |
| 55 | PACIF | Pacific | 7,001 | 2 | Time Based | 1.24 |
| 56 | LIBTY | Liberty | 35,300 | 5 | Time Based | 4.60 |
| 57 | KCWST | Kansas City | 50,000 | 22.5 | Time Based | 18.13 |

^a^ Unit: million gallon per day (MGD).

^b^ Average flow: calculated for the period July 6, 2020, to December 7, 2020.

**Table S2.** The coefficients, standard error, t value and P value in Forward and backward stepwise regression models (Positive ion mode)

| Chemicals | Coefficients | Standard Error | t value | ^a^ Pr (>\|t\|) |
| --- | --- | --- | --- | --- |
| (Intercept) | -3.273e+01 | 8.510e+00 | -3.846 | 0.06144 |
| Netilmicin | -2.299e-03 | 2.975e-04 | -7.727 | 0.01634 * |
| Trolamine | -1.679e-03 | 1.687e-04 | -9.957 | 0.00994 ** |
| 3-Chloro-4-(dichloromethyl)-5-hydroxy-2(5H)-furanone | 7.009e-05 | 1.093e-05 | 6.414 | 0.02346 * |
| Dimethicone | 1.189e-02 | 2.133e-03 | 5.574 | 0.03071 * |
| 4-Dodecylphenol | -8.137e-06 | 1.511e-06 | -5.386 | 0.03278 * |
| 2-Dodecylbenzenesulfonic acid | -1.390e-03 | 1.196e-04 | -11.621 | 0.00732 ** |
| Cetrimonium | 4.775e-04 | 7.426e-05 | 6.430 | 0.02335 * |
| Diethylene glycol | -1.520e-03 | 1.246e-03 | -1.220 | 0.34681 |
| 1-Octadecanamine | -1.242e-05 | 1.093e-04 | -0.114 | 0.91995 |
| Aluminium dodecanoate | 4.199e-03 | 5.266e-04 | 7.974 | 0.01536 * |
| 2-Diethylaminoethanol | -6.275e-04 | 1.427e-04 | -4.397 | 0.04803 * |
| Palmitelaidic acid | 3.682e-03 | 3.485e-04 | 10.565 | 0.00884 ** |
| 4-Nonylphenol | 5.423e-03 | 8.503e-04 | 6.378 | 0.02371 * |
| Dicyclopentadiene | 5.268e-04 | 1.407e-04 | 3.743 | 0.06454 . |
| Nonoxynol-9 | 1.581e-03 | 2.342e-04 | 6.750 | 0.02125 * |
| N-Undecylbenzenesulfonic acid | 2.173e-03 | 3.280e-04 | 6.625 | 0.02204 * |
| Dicyclohexylamine | 2.441e-03 | 2.330e-04 | 10.474 | 0.00899 ** |
| Tetrabutylammonium | 1.119e-03 | 8.918e-05 | 12.553 | 0.00629 ** |
| Sodium Tetradecyl Sulfate | -5.511e-03 | 6.154e-04 | -8.956 | 0.01224 * |
| Sodium oleate | 4.822e-02 | 5.683e-03 | 8.485 | 0.01361 * |
| Cetrimide | 1.100e-02 | 1.050e-03 | 10.481 | 0.00898 ** |

^a^ Significance codes: 0 '***' 0.001 '**' 0.01 '*' 0.05 '.' 0.1 ' ' 1

**Table S3.** The coefficients, standard error, t value and P value in best subset regression model (Positive ion mode)

| **Chemicals** | **Coefficients** | **Standard Error** | **t value** | **^a^ Pr (>\|t\|)** |
| --- | --- | --- | --- | --- |
| ## (Intercept) | -3.314e+01 | 6.331e+00 | -5.234 | 0.013572 * |
| Netilmicin | -2.327e-03 | 1.407e-04 | -16.534 | 0.000482 *** |
| Trolamine | -1.693e-03 | 1.014e-04 | -16.696 | 0.000468 *** |
| 3-Chloro-4-(dichloromethyl)-5-hydroxy-2(5H)-furanone | 7.086e-05 | 7.034e-06 | 10.073 | 0.002084 ** |
| Dimethicone | 1.205e-02 | 1.291e-03 | 9.333 | 0.002605 ** |
| 4-Dodecylphenol | -8.117e-06 | 1.229e-06 | -6.604 | 0.007069 ** |
| 2-Dodecylbenzenesulfonic acid | -1.388e-03 | 9.680e-05 | -14.335 | 0.000736 *** |
| Cetrimonium | 4.830e-04 | 4.589e-05 | 10.524 | 0.001832 ** |
| Diethylene glycol | -1.635e-03 | 5.951e-04 | -2.748 | 0.070866 . |
| Aluminium dodecanoate | 4.241e-03 | 3.100e-04 | 13.678 | 0.000845 *** |
| 2-Diethylaminoethanol | -6.402e-04 | 7.298e-05 | -8.772 | 0.003121 ** |
| Palmitelaidic acid | 3.711e-03 | 1.889e-04 | 19.644 | 0.000288 *** |
| 4-Nonylphenol | 5.490e-03 | 5.042e-04 | 10.889 | 0.001658 ** |
| Dicyclopentadiene | 5.191e-04 | 1.009e-04 | 5.144 | 0.014234 * |
| Nonoxynol-9 | 1.601e-03 | 1.249e-04 | 12.812 | 0.001026 ** |
| N-Undecylbenzenesulfonic acid | 2.198e-03 | 2.017e-04 | 10.896 | 0.001654 ** |
| Dicyclohexylamine | 2.463e-03 | 1.029e-04 | 23.943 | 0.000160 *** |
| Tetrabutylammonium | 1.124e-03 | 6.440e-05 | 17.457 | 0.000410 *** |
| Sodium Tetradecyl Sulfate | -5.566e-03 | 3.128e-04 | -17.794 | 0.000387 *** |
| Sodium oleate | 4.872e-02 | 2.950e-03 | 16.514 | 0.000483 *** |
| Cetrimide | 1.108e-02 | 6.711e-04 | 16.506 | 0.000484 *** |

^a^ Significance codes: 0 '***' 0.001 '**' 0.01 '*' 0.05 '.' 0.1 ' ' 1

**Table S4.** The coefficients in lasso regression model (Positive ion mode)

| Chemicals | Coefficients |
| --- | --- |
| (Intercept) | 4.686844e+01 |
| Netilmicin | -2.370461e-05 |
| Trolamine | 0.000000e+00 |
| 3-Chloro-4-(dichloromethyl)-5-hydroxy-2(5H)-furanone | -3.751467e-05 |
| Dimethicone | -1.038730e-02 |
| 4-Dodecylphenol | -6.466841e-12 |
| 2-Dodecylbenzenesulfonic acid | -6.102094e-04 |
| Cetrimonium | 0.000000e+00 |
| Diethylene glycol | 5.582177e-03 |
| 1-Octadecanamine | -7.685889e-04 |
| Aluminium dodecanoate | -2.879951e-04 |
| 2-Diethylaminoethanol | 3.874359e-04 |
| Palmitelaidic acid | 8.952359e-04 |
| 4-Nonylphenol | 8.041543e-04 |
| Dicyclopentadiene | 2.984327e-05 |
| Nonoxynol-9 | 0.000000e+00 |
| Stearic acid | 0.000000e+00 |
| N-Undecylbenzenesulfonic acid | 0.000000e+00 |
| Dicyclohexylamine | 0.000000e+00 |
| Tetrabutylammonium | 8.733960e-05 |
| Sodium Tetradecyl Sulfate | 3.165432e-04 |
| Sodium oleate | 4.652897e-03 |

**Table S5.** The coefficients in lasso regression model (Negative ion mode)

| Chemicals | Coefficients |
| --- | --- |
| (Intercept) | 10.8395801799 |
| 2-Dodecylbenzenesulfonic acid | 0.0000000000 |
| 4-Octylphenol | 0.0000000000 |
| Polyoxyethylene dioleate | 0.0000000000 |
| Sodium Tetradecyl Sulfate | 0.0004798996 |
| 4-Nonylphenol | 0.0006729771 |
| Oleic Acid | -0.0010412793 |
| Lauroyl peroxide | 0.0000000000 |
| Palmitic acid | 0.0000000000 |

**Table S6.** The coefficients, standard error, t value and P value in forward stepwise regression model (Negative ion mode)

| **Chemicals** | **Coefficients** | **Standard Error** | **t value** | **^a^ Pr (>\|t\|)** |
| --- | --- | --- | --- | --- |
| (Intercept) | 8.6956453 | 5.9985551 | 1.450 | 0.1627 |
| Sodium tetradecyl sulfate | 0.0005345 | 0.0000903 | 5.920 | 8.64e-06 *** |
| 4-Nonylphenol | 0.0008837 | 0.0003412 | 2.590 | 0.0175 * |
| Oleic Acid | -0.0014011 | 0.0006643 | -2.109 | 0.0477 * |

^a^Significance codes: 0 '***' 0.001 '**' 0.01 '*' 0.05 '.' 0.1 ' '
